# Implications of *Dobbs v.* Jackson for patients and providers: a scoping review

**DOI:** 10.1101/2023.07.10.23292460

**Authors:** David T. Zhu, Lucy Zhao, Tala Alzoubi, Novera Shenin, Teerkasha Baskaran, Julia Tikhonov, Catherine Wang

## Abstract

**Introduction:** On June 24, 2022, the U.S. Supreme Court’s decision in *Dobbs v. Jackson* overturned the right to abortion set forth by *Roe v. Wade*, granting states the authority to regulate access to abortion services. This has led to widespread bans, threatening patients’ access to, and healthcare providers’ abilities to provide, the full spectrum of reproductive health services. The ruling disproportionately affects marginalized groups, exacerbating existing social disparities in health and is an emerging public health crisis.

**Methods:** We conducted a scoping review to evaluate the impact of *Dobbs* on patients’ health outcomes and access to health services, as well as on medical trainees’ and healthcare providers’ ability to access abortion training and provide reproductive health services. The search was based on the PRISMA Extension for Scoping Reviews (PRSIMA-ScR) guidelines. We searched eight bibliographic databases (PubMed, Scopus, Embase, PsycINFO, Google Scholar, Science Direct, JSTOR, and Web of Science) and three preprint servers (medRxiv, bioRxiv, and Europe PMC) using various combinations of keywords related to ‘abortion’ and ‘Dobbs v. Jackson’ on March 22, 2023. Four reviewers independently screened the studies based on pre-specified eligibility criteria and one reviewer performed data extraction for pre-identified themes.

**Results:** A total of 18 studies met the inclusion criteria. We found that *Dobbs* led to a surge in demand for contraception, compounded existing travel- and cost-related barriers to access, increased polarizing views on social media (e.g., Twitter), and evoked significant fears and concerns among medical trainees regarding their scope of practice and fears of legal repercussions for offering standard-of-care and related services to patients seeking abortions.

**Conclusion:** Our study offers valuable insights into the clinical implications of *Dobbs* on patients’ health outcomes and access to health services, as well as providers’ reproductive health practices.

## 1. Introduction

The U.S. Supreme Court’s decision in *Dobbs v. Jackson Women’s Health Organization* in June 2022 overturned nearly a half-century of the constitutionally protected right to abortion in the U.S. initially set forth by *Roe v. Wade* in 1973.^1, 2^ This ruling grants individual states the authority to regulate access to abortion care and reproductive health services with varying degrees of restrictions.^1, 2^ To date, approximately half of the U.S. states have imposed partial or complete bans on the provision of induced abortions.^2, 3^ Abortions are both safe and effective, and there is extensive global evidence that restricting access to legal abortions does not lower the overall rate of abortions; rather, it merely limits the rate of legal abortions, thus, increasing the rate of unsafe abortions, which are more prone to adverse health outcomes and complications.^4, 5^

The implications of *Dobbs v. Jackson* on patients’ health outcomes and providers’ ability to provide the full spectrum of reproductive health services have been harmful and widespread. Roughly 60% of women of reproductive age live in U.S. states that are “hostile” to abortions.^3^ A recent study predicts that a nationwide abortion ban would increase maternal mortality from childbirth or pregnancy complications by 21% in the general U.S. population and 33% among Black Americans.^6^ *Dobbs* is likely to stretch existing social and reproductive health disparities, given that abortions are disproportionately needed by patients from low-income backgrounds, those who identify with one or more racial/ethnic minorities, and other medically-underserved groups.^7–9^ *Dobbs* is also likely to exacerbate existing barriers to access abortion care, including cost and travel; e.g., those living in abortion-hostile states will likely need to travel out-of-state to access abortion or reproductive health services.^10^ The previous literature shows that women who are unable to terminate a pregnancy against their wishes were more likely to experience continued intimate partner violence (IPV) compared to those who had access to comprehensive services for pregnancy termination.^11^ This may also have collateral effects on other fields, such as healthcare provision for congenital diseases and neonates.^12^

To the best of our knowledge, there have been no other reviews that have comprehensively evaluated the clinical implications of *Dobbs* on patients and providers, likely due to the novelty of this health issue. To address this gap in the literature, we conducted a scoping review to overview the impact of *Dobbs* on (1) patients’ health outcomes or access to abortions and reproductive health services; (2) medical trainees’ access to comprehensive abortion training; and (3) providers’ ability to provide the full spectrum of reproductive health services.

## 2. Methods

### 2.1 Methodological approach

We conducted a scoping review in accordance with the methodological framework created by Arksey & O’Malley^13^ and the PRISMA extension for Scoping Reviews (PRISMA-ScR) guidelines^14^ **(Table S1)** to capture both peer-reviewed and grey literature related to the clinical implications of Dobbs v. Jackson. Given the novelty and rapidly evolving nature of this topic, the flexibility and breadth of a scoping review is well-suited to address our research objectives.

### 2.2 Information sources and search strategy

We conducted a literature search in eight bibliographic databases (PubMed, Scopus, Embase, PsycINFO, Google Scholar, Science Direct, JSTOR, and Web of Science) to capture published peer-reviewed studies, in addition to three other servers (medRxiv, bioRxiv, and Europe PMC) to capture preprint studies. Various combinations of the search terms “Dobbs”, “Roe”, “abortion”, “pregnancy termination”, “unintended pregnancy”, “abortifacient”, “misopristol”, “mifeprex”, “mifepristone”, “cytotec”, were used to retrieve articles on March 22, 2023. Given that the *Dobbs v. Jackson* ruling occurred in June 2022, we restricted our search from 2022 to 2023 to optimize our search. The detailed search strategy, including combinations of MeSH terms and Boolean operators, can be found in **(Table S2)**.

### 2.3 Selection of sources of evidence

Four reviewers (LZ, TA, NS, TB) independently screened the title and abstracts of retrieved articles based on pre-specified inclusion and exclusion criteria **(Table 1)**. Abstracts were included. At this stage, studies that were classified as being potentially relevant subsequently underwent full-text screening by the same reviewers. The final subset of studies included in this review were verified and approved by all reviewers. Any screening conflicts that arose were resolve by a neutral fifth reviewer (DTZ).

**Table 1.** Eligibility criteria.

|  |
| --- |
| **The following articles were included:** |
| Any adult patient population (either general or specialized) based in the United States |
| Includes the exposure of interest (i.e., *Dobbs v. Jackson* ruling) |
| Evaluates the patient-centered outcomes of interest (i.e., access to abortion and reproductive health services, health outcomes related to physical and psychological wellbeing) |
| Evaluates the provider-centered outcomes of interest (i.e., access to comprehensive abortion training, facilitators and barriers to providing abortion services) |
| English language |
| Studies with primary quantitative data (e.g., primary observational studies) |
| Published after May 2, 2022 (when the U.S. Supreme Court’s draft of *Dobbs* became leaked) |
| **The following articles were excluded:** |
| Patient populations outside the United States |
| Does not include the exposure of interest |
| Does not include information on the patient- or provider-centered outcomes of interest |
| No English language |
| Studies with only qualitative data |
| Studies with only secondary data (e.g., reviews, editorials, letters to the editor, etc.) |
| Published before May 2, 2022 |

### 2.4 Data charting process and items

Following full-text screening, one reviewer (DTZ) performed data extraction and all other co-authors verified the data. Relevant information was collected and inputted into a data extraction form with prespecified endpoints such as the publication year, authors, study design, data collection period, data sources, methods and data analysis, and the main outcomes and findings related to abortion and reproductive health services. The data extraction template consisted of two sections (one for patient-oriented studies and another for studies involving medical trainees and healthcare providers). Preprints and abstracts remaining after screening were updated with their final peer-reviewed versions if available.

### 2.5 Analysis, synthesis, and presentation of results

The final sample of studies were analyzed thematically, such as discussion of contraception [e.g., permanent contraception (PC), emergency contraception (EC), and other forms of contraception], facilitators and barriers that patients (and providers) face with accessing (and providing) care after *Dobbs*, health outcomes, public attitudes, and other relevant themes to abortion and reproductive health services in the post-*Dobbs* landscape.

## 3. Results

### 3.1 Sample and article characteristics

Our initial search yielded 2,609 articles. Through automatic deduplication by Covidence, 936 articles were removed. Title and abstract screening removed another 1,638 articles and, lastly, full-text screening removed an additional 17 articles. Our final sample consisted of 18 articles (i.e., 12 full-text articles and 6 abstracts) which all underwent data extraction. Our screening process yielded a Cohen’s Kappa score of 0.82. An overview of our screening process is presented in **(Figure 1)**. The study designs were predominantly cross-sectional (n=6), modeling (n=5), and observational (n=4), and additionally, there was one retrospective chart review, one NLP-based study, and one commentary **(Table 2)**.

**Table 2.**
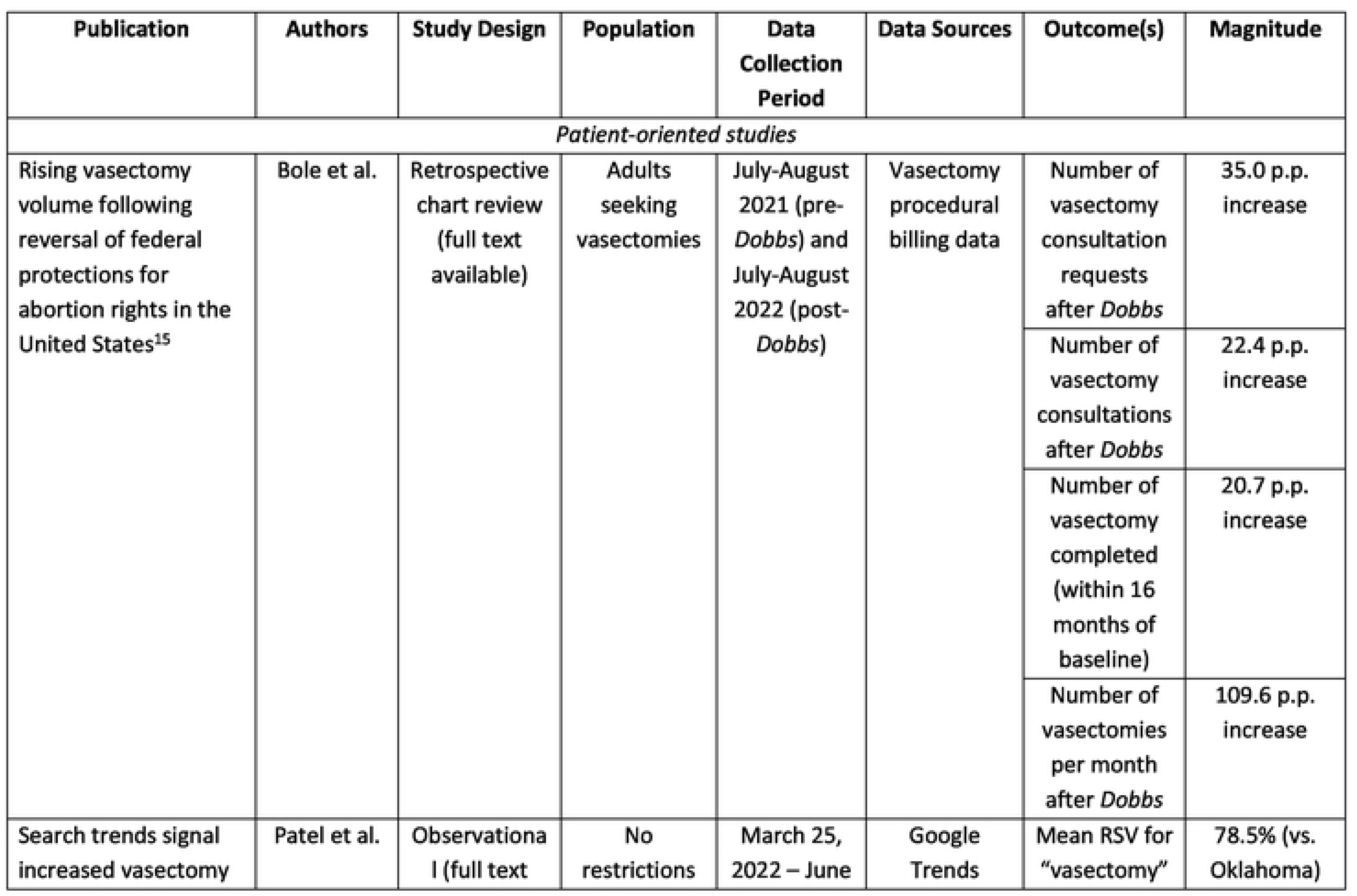
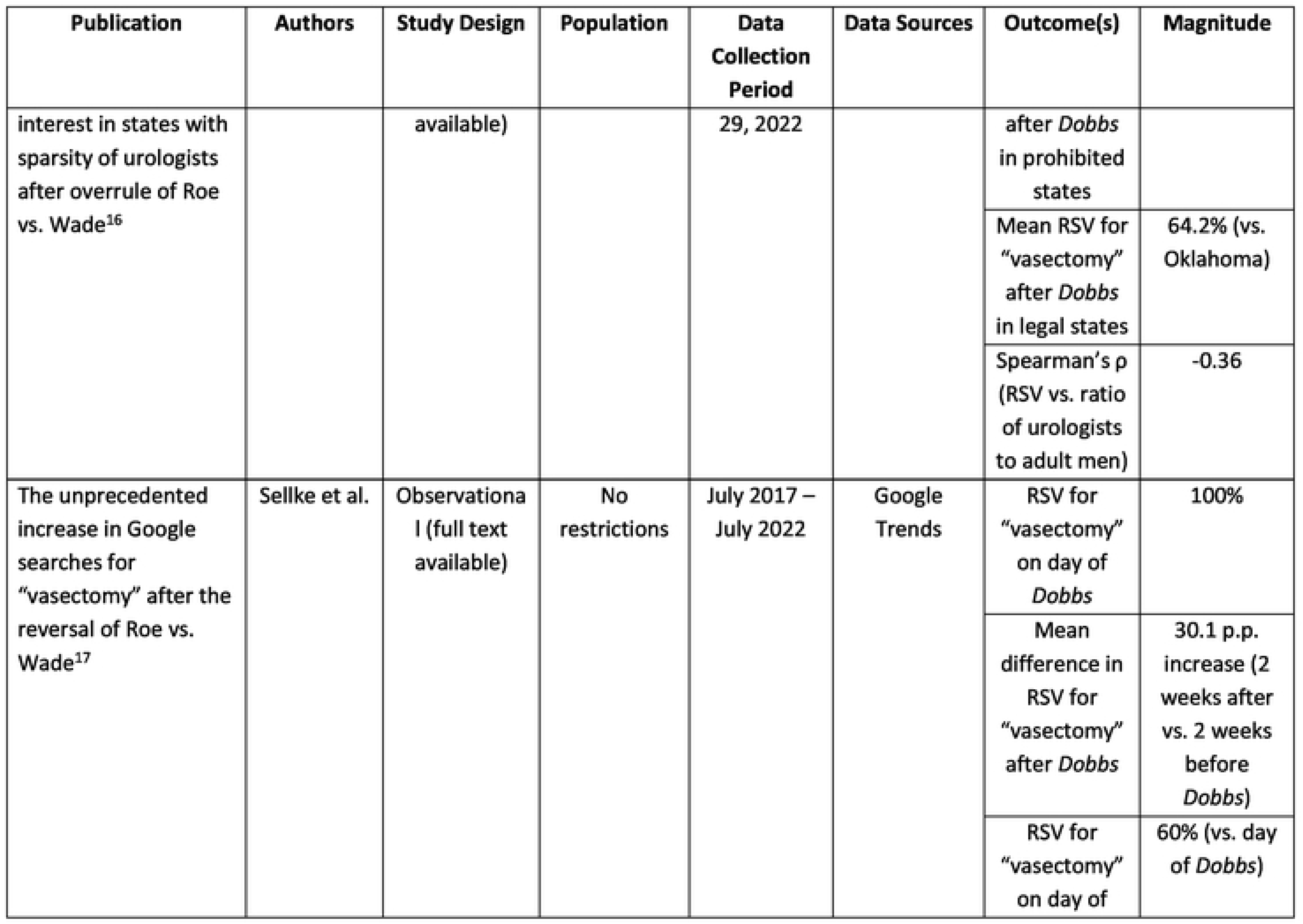
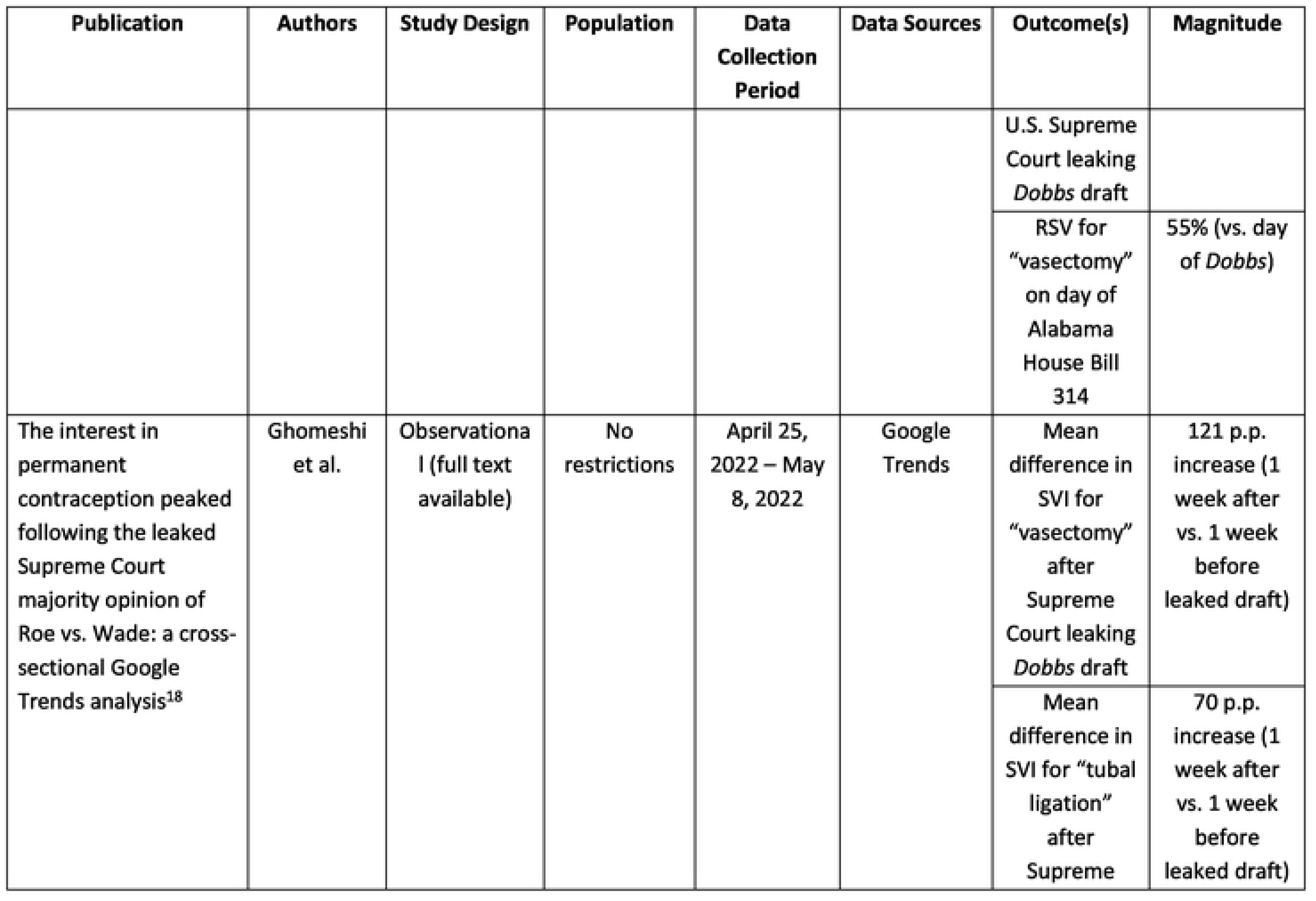
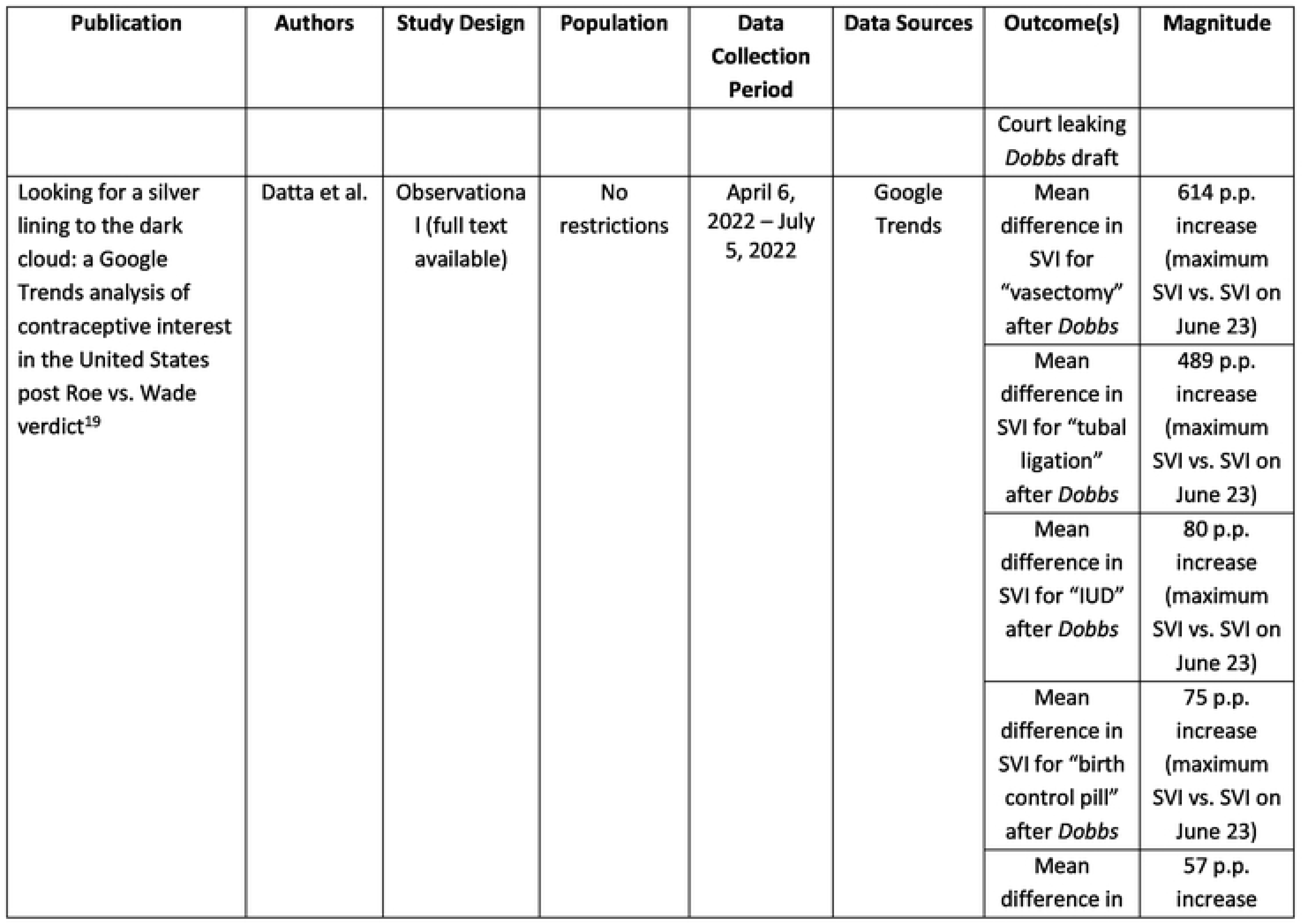
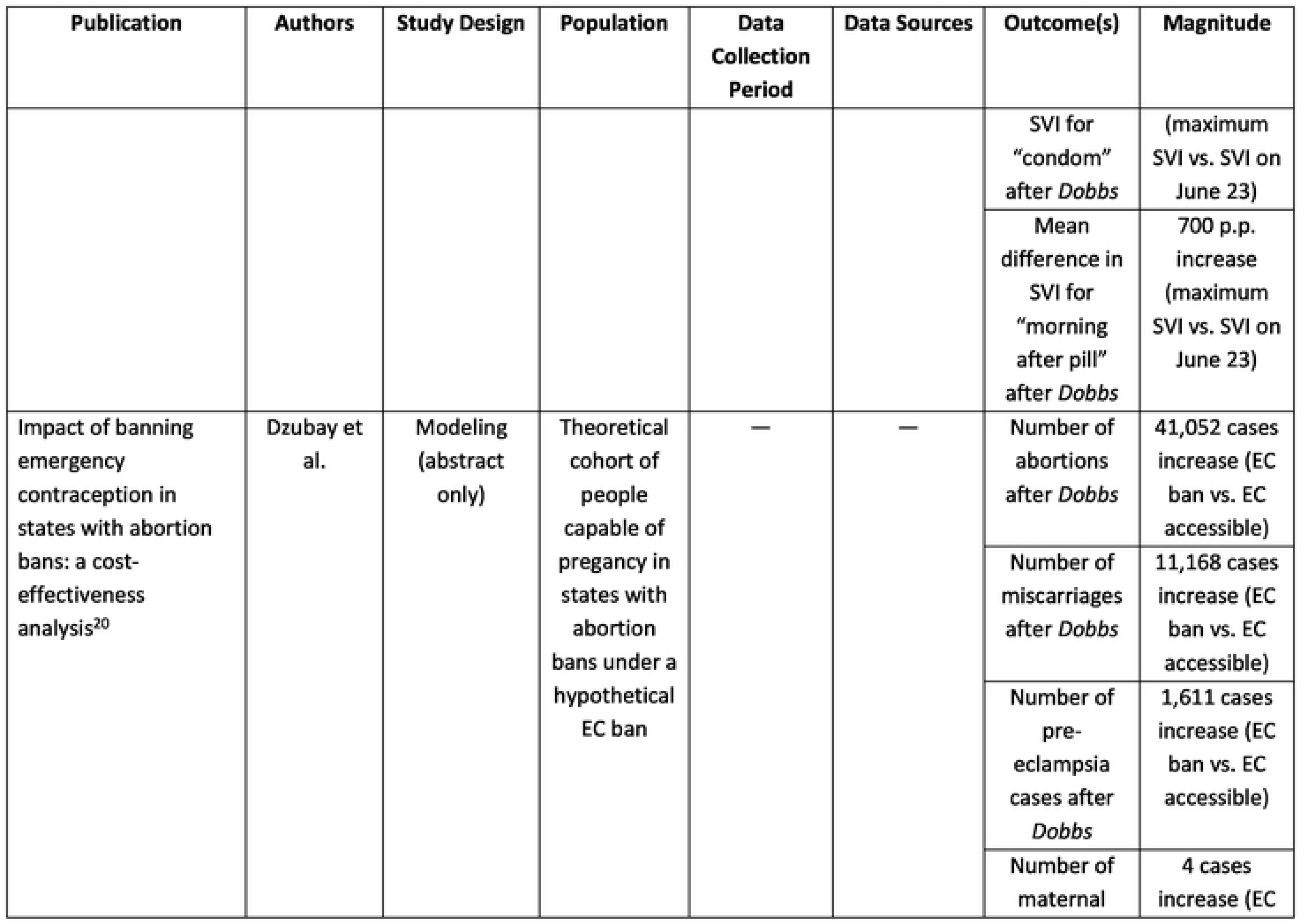
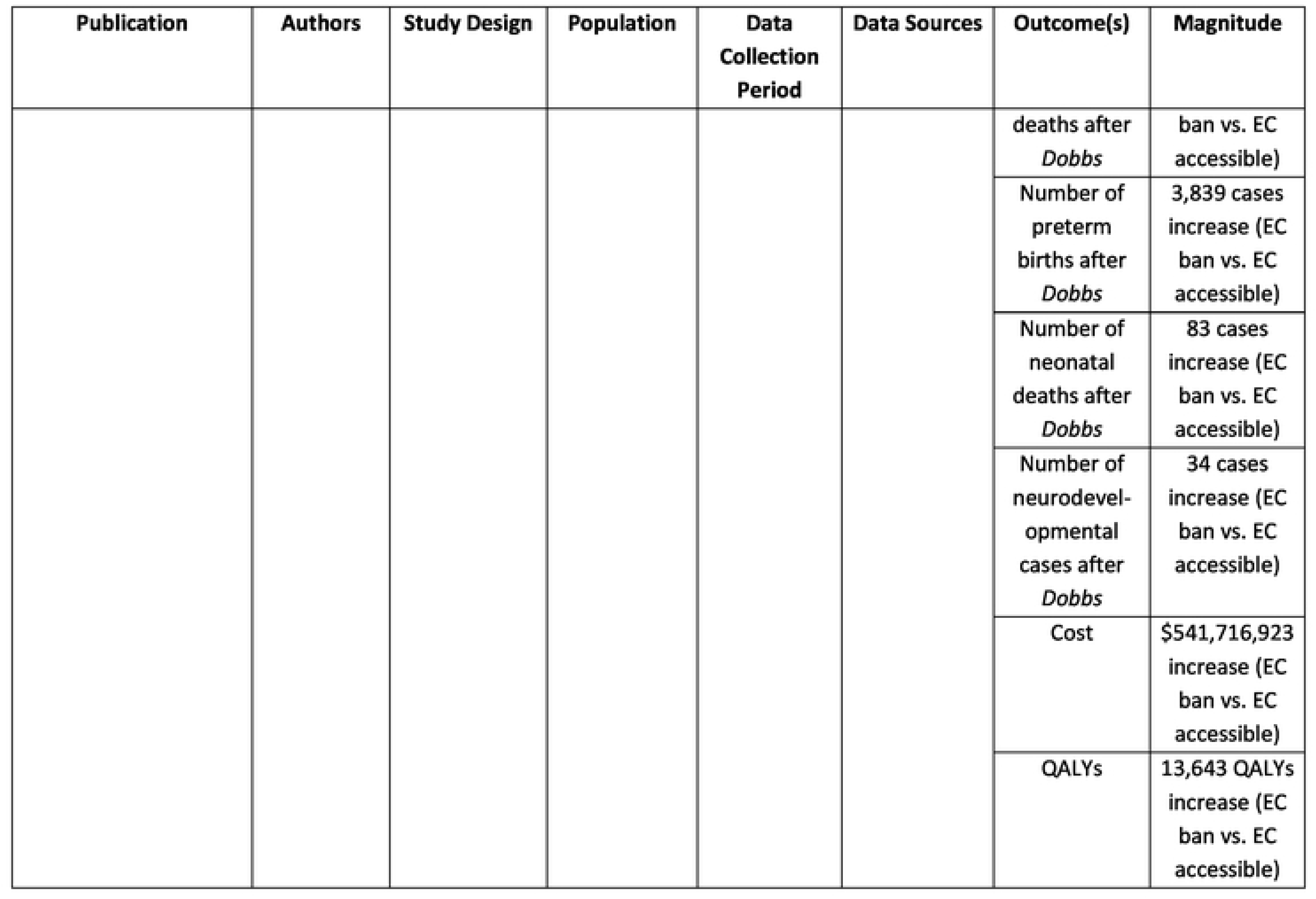
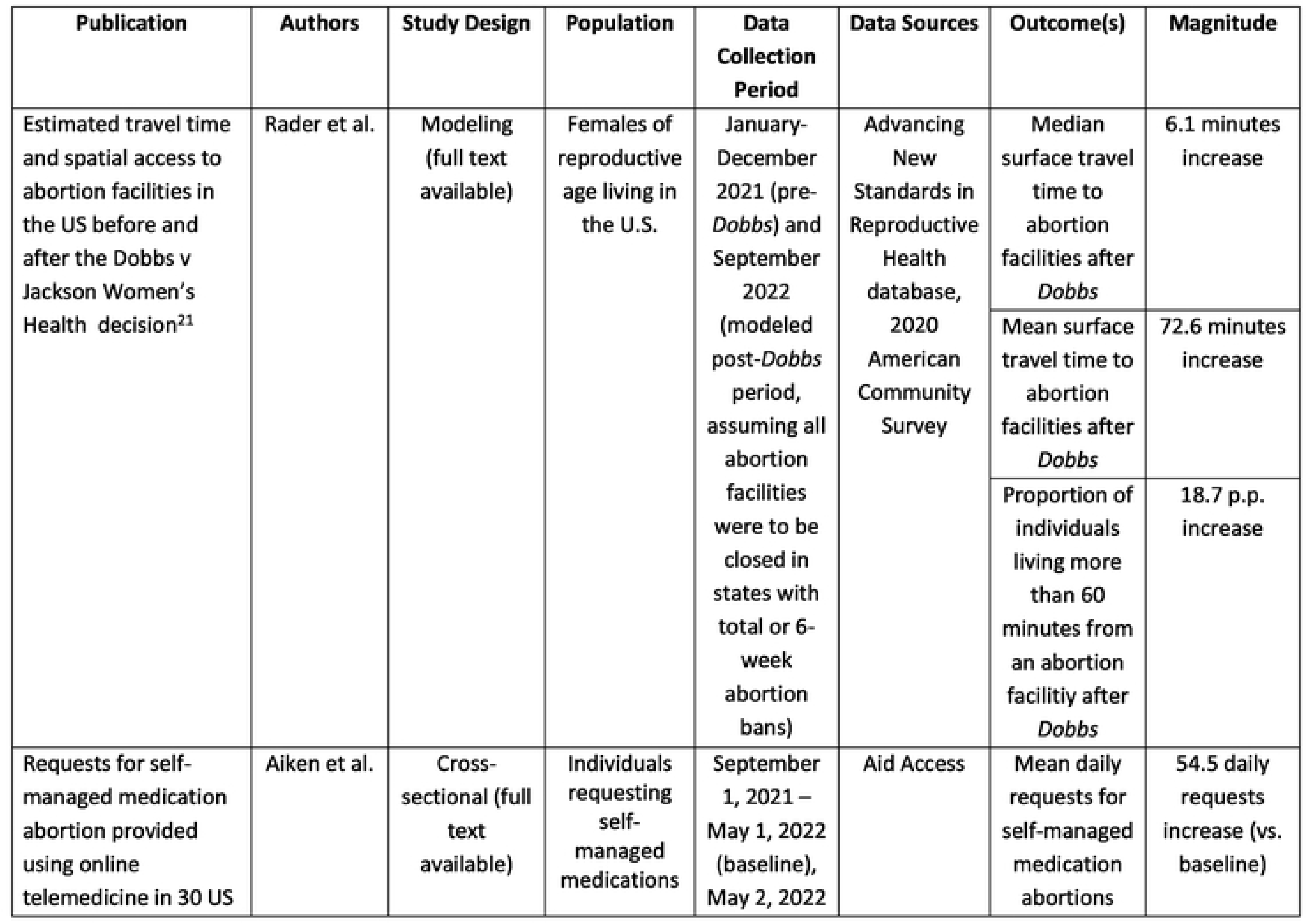
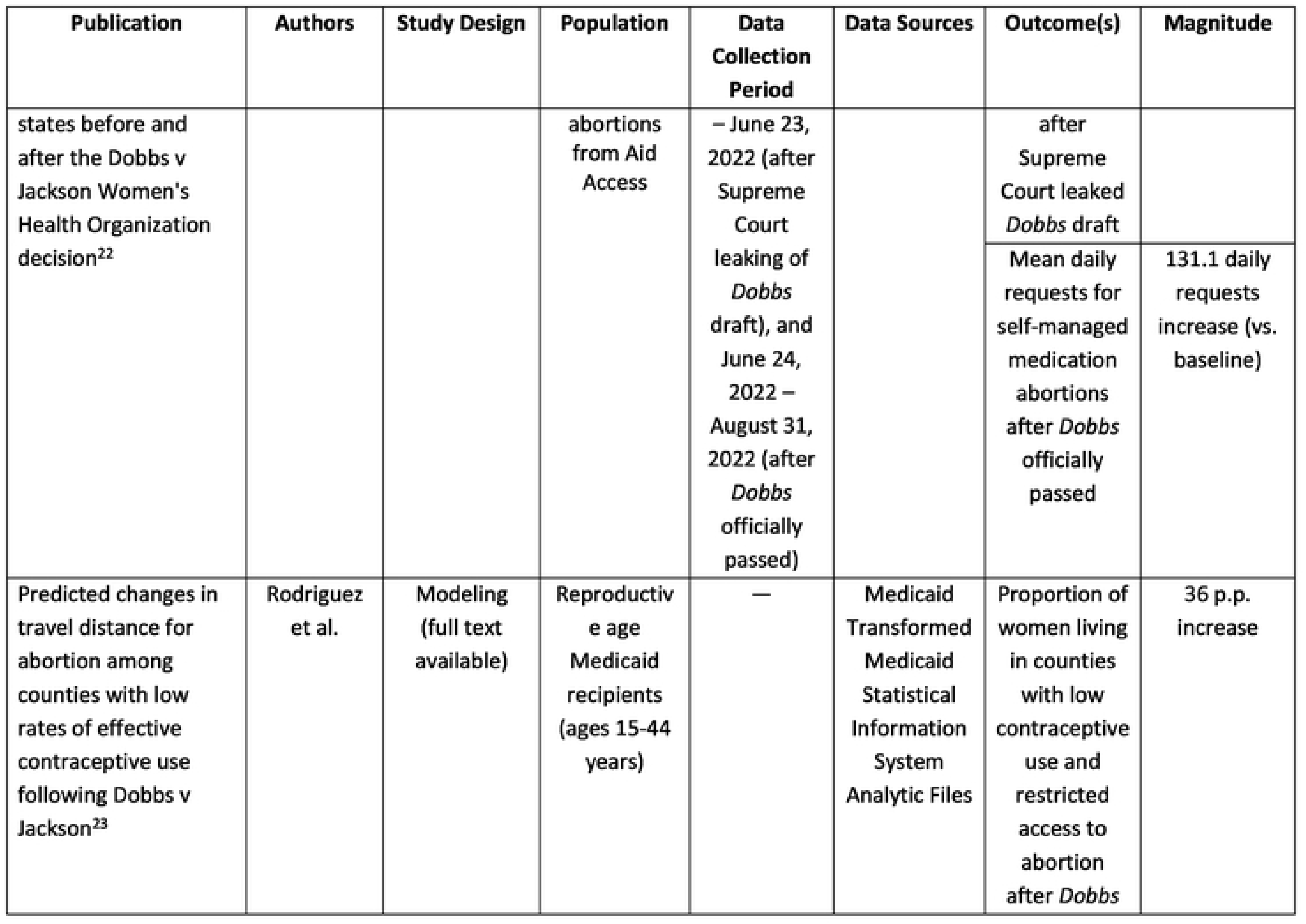
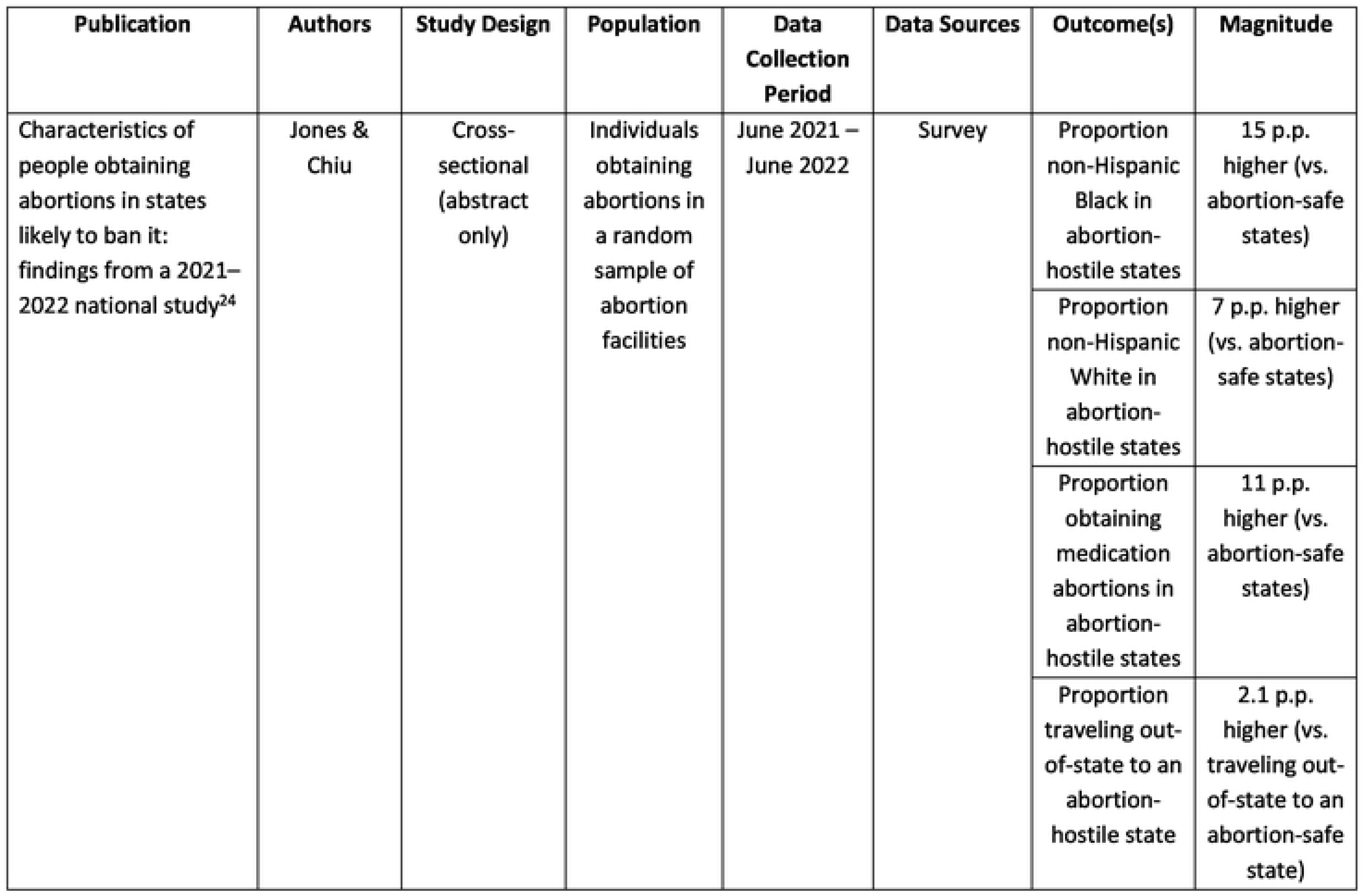
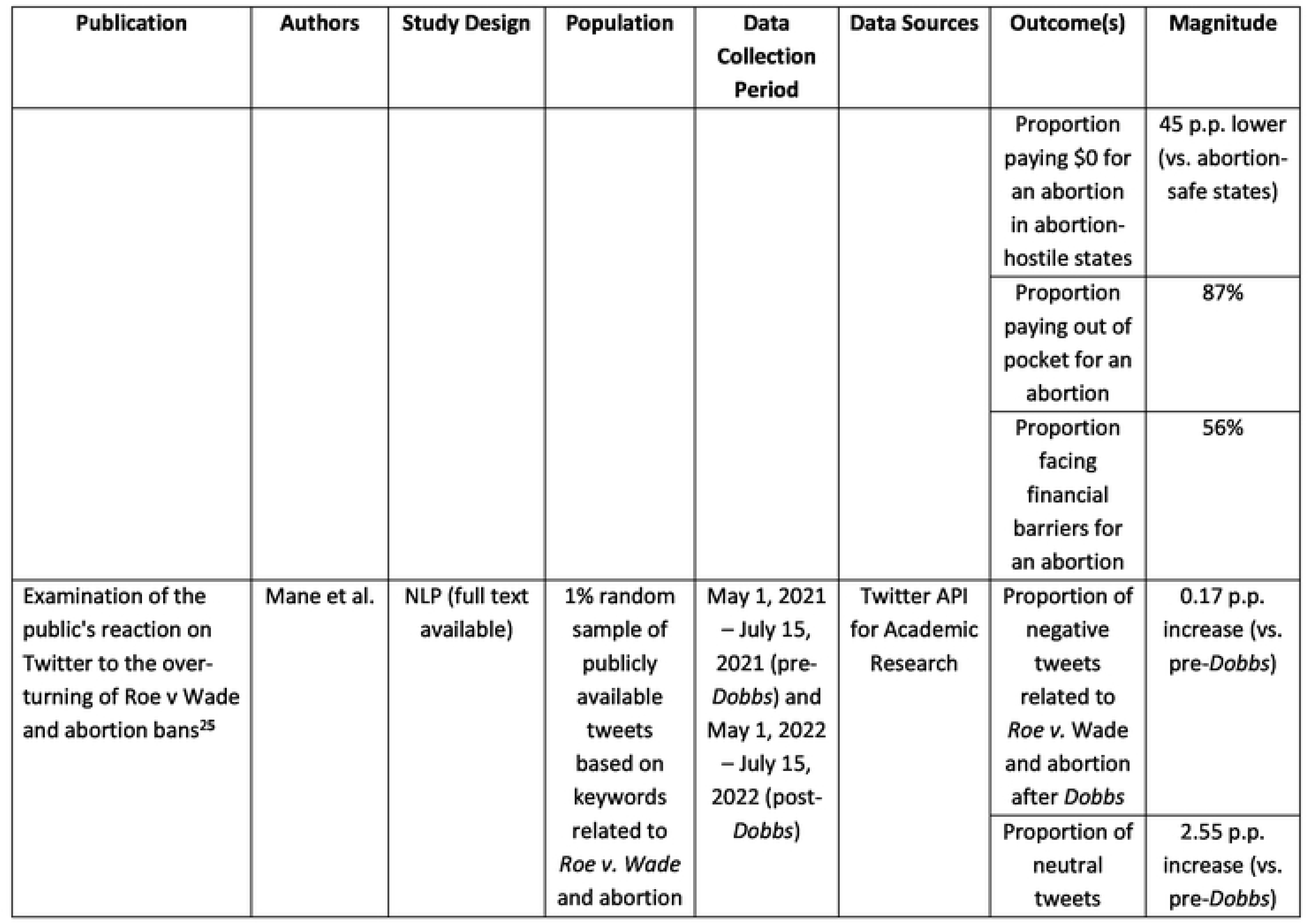
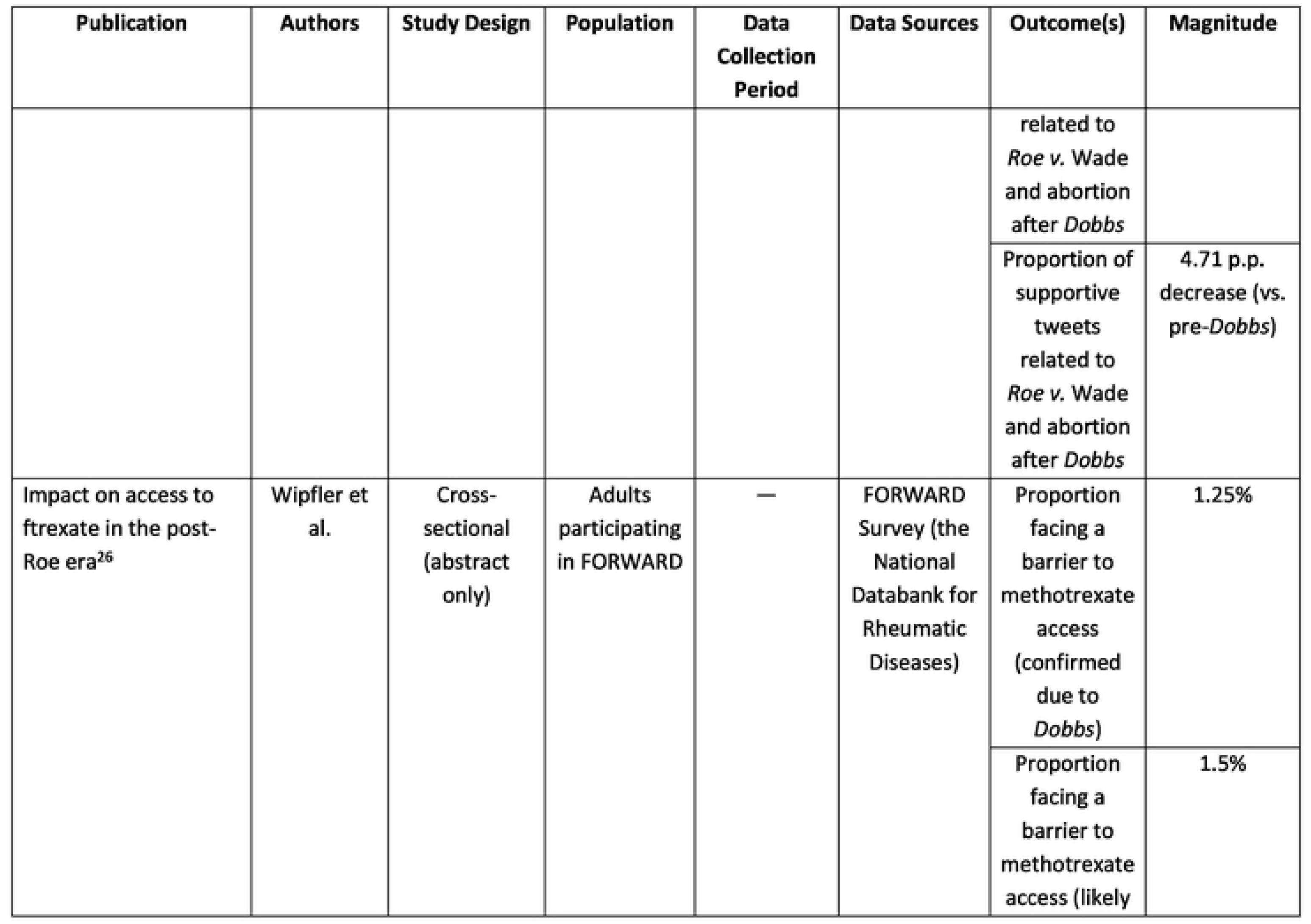
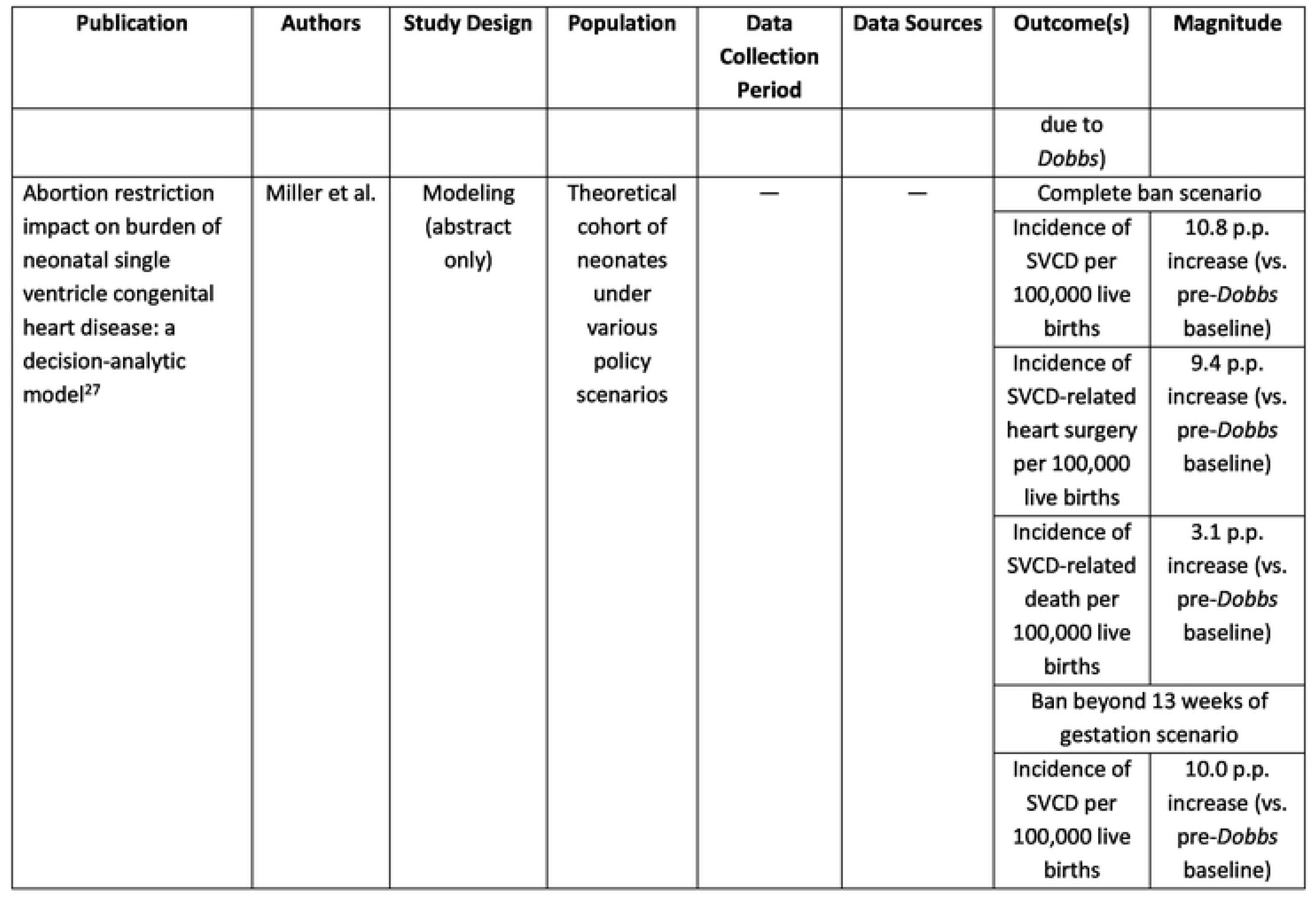
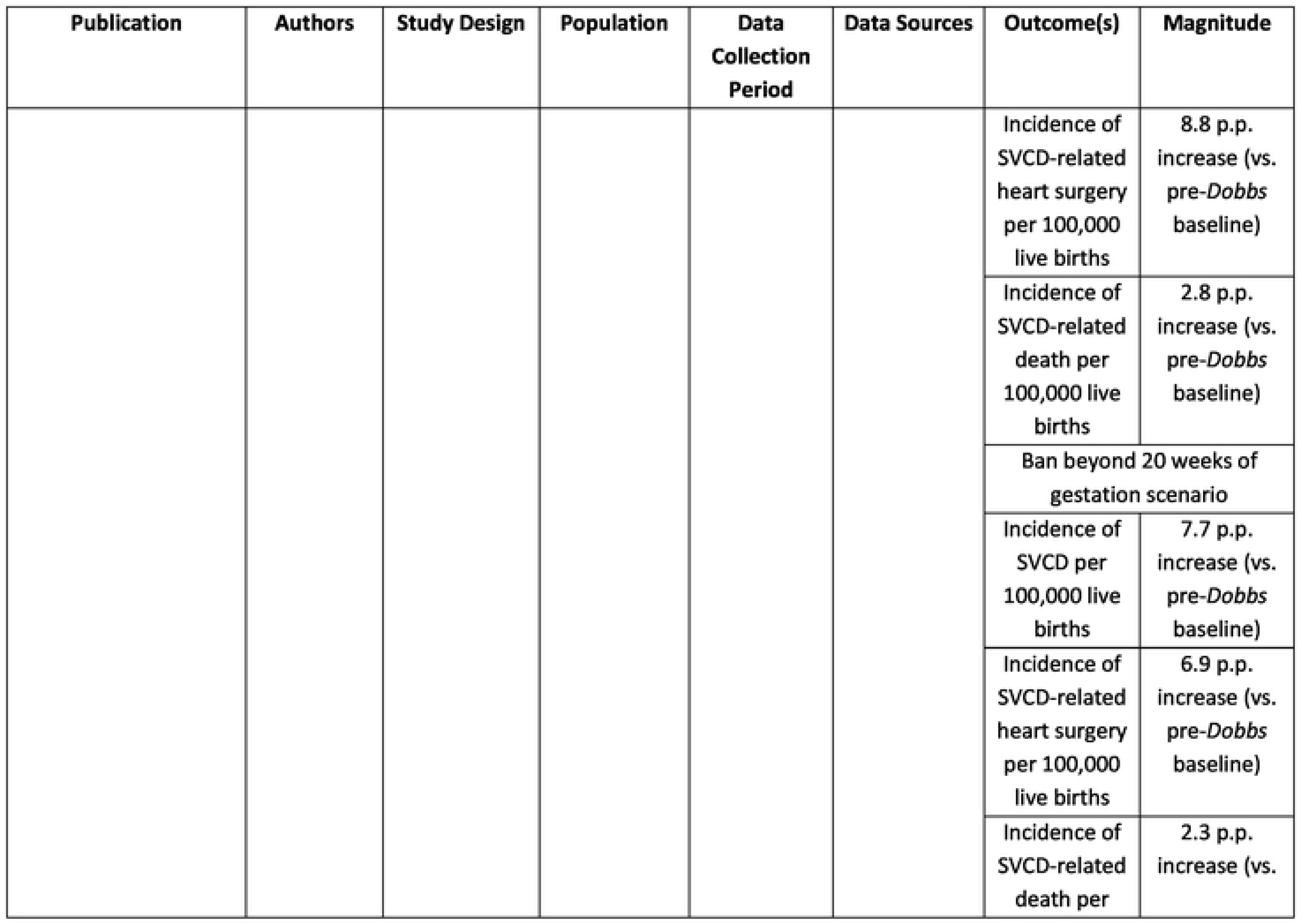
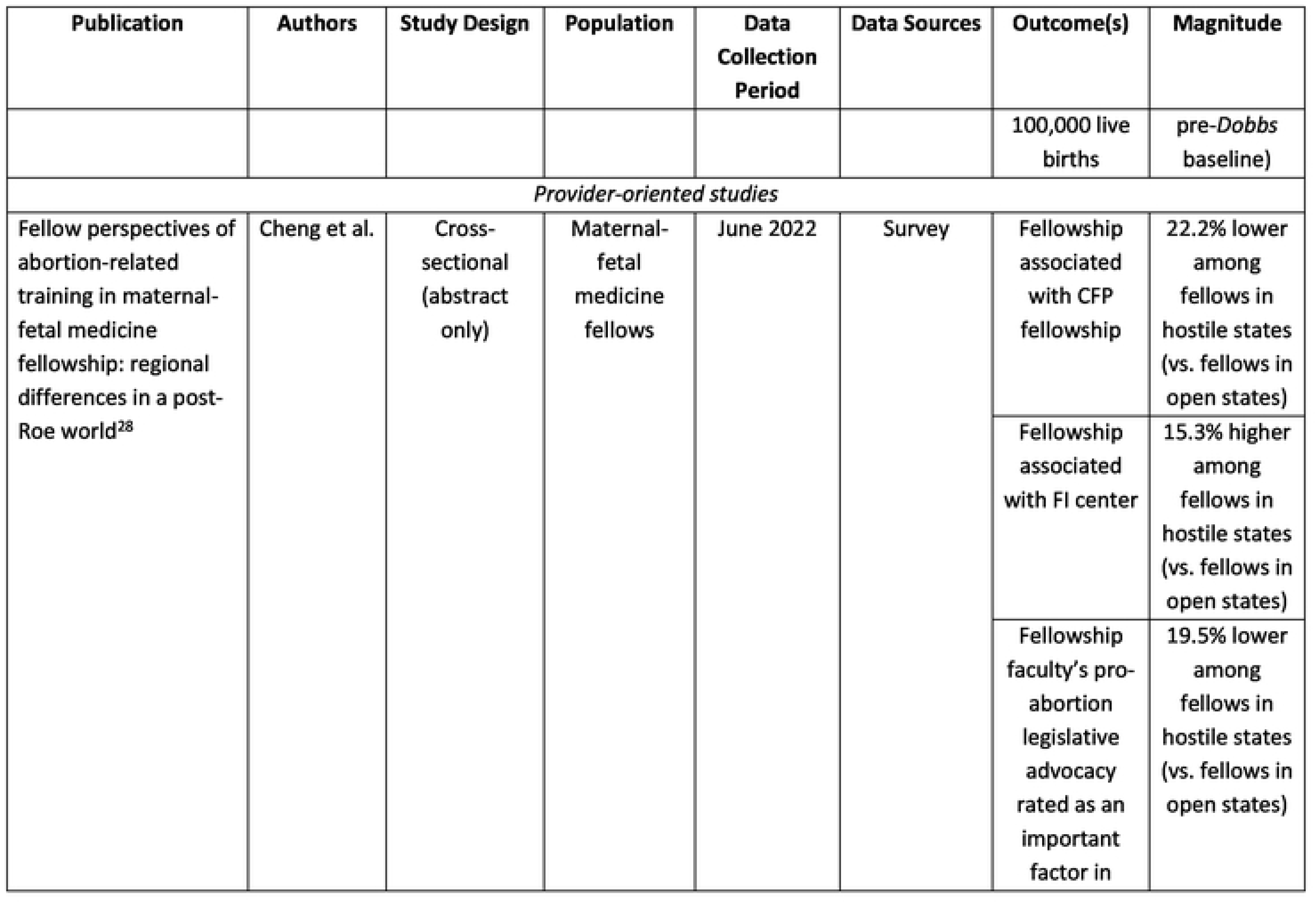
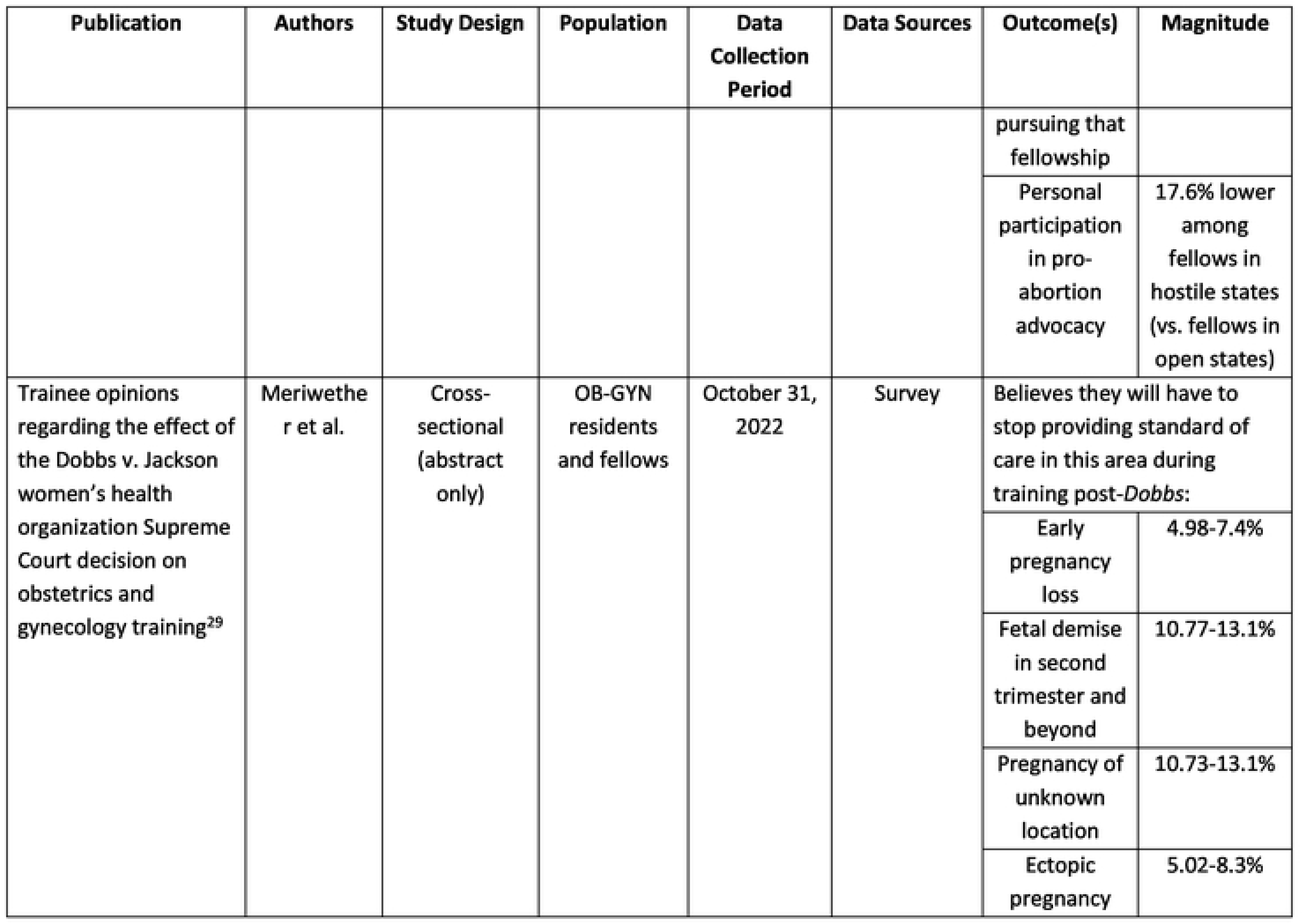
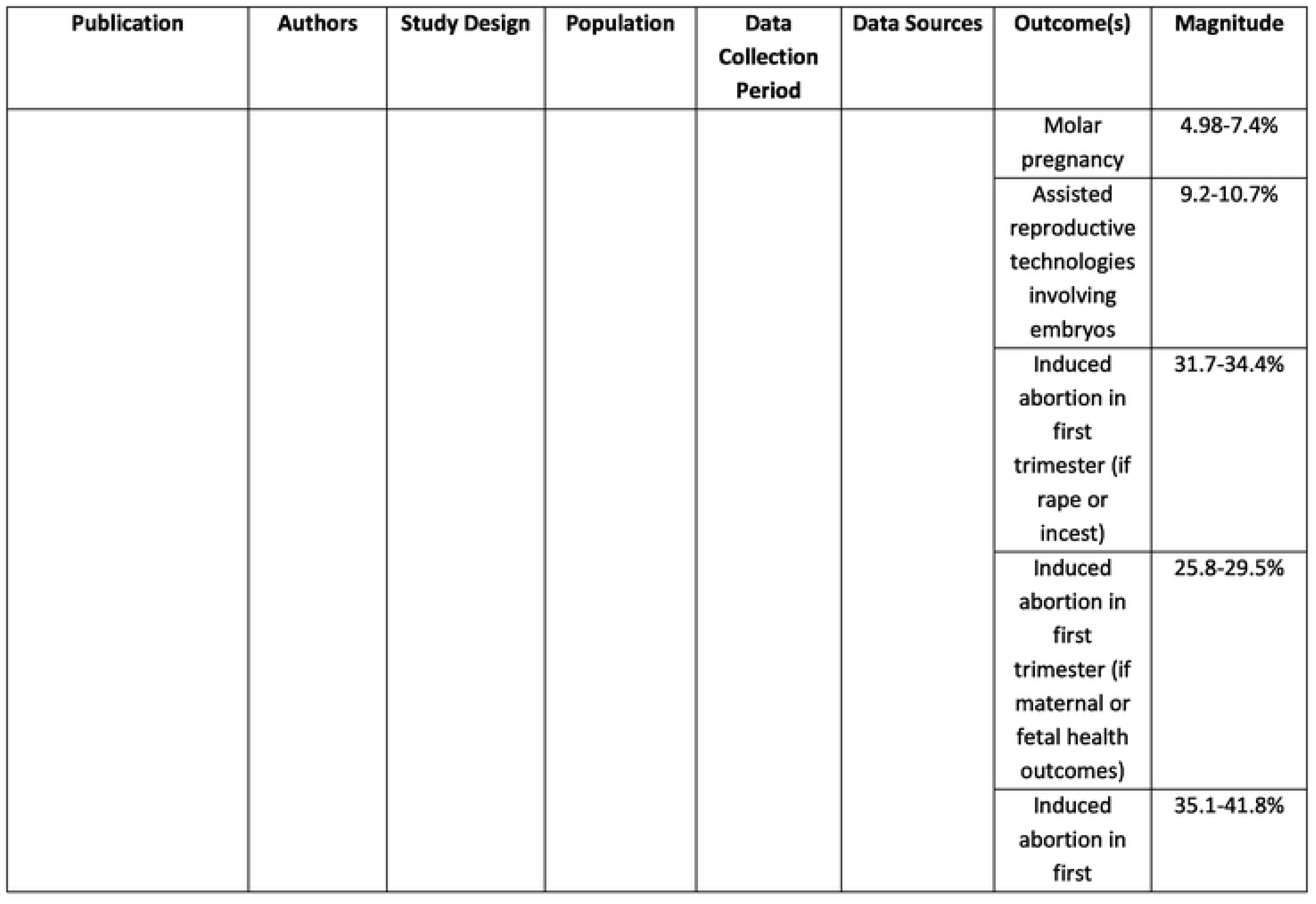
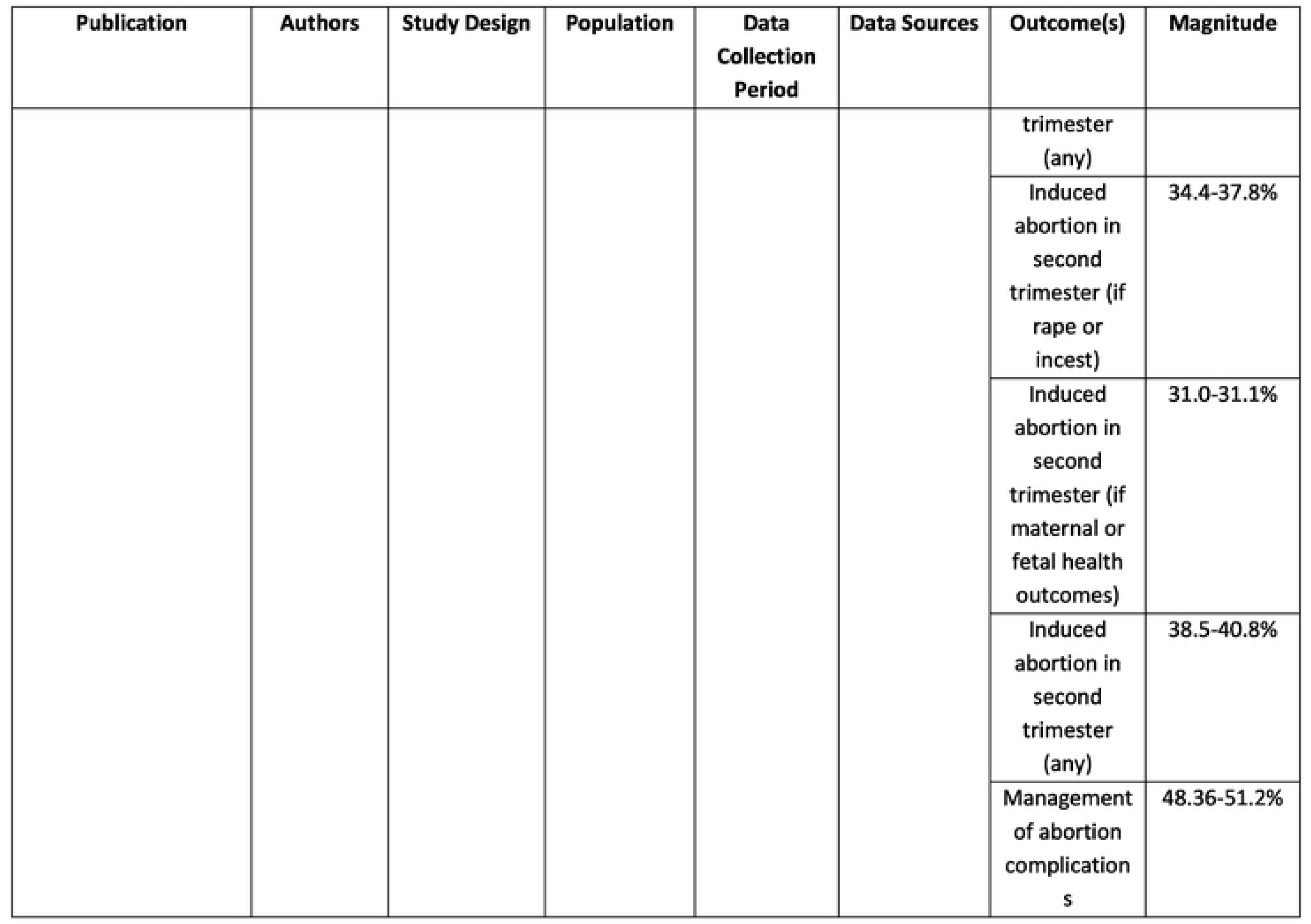
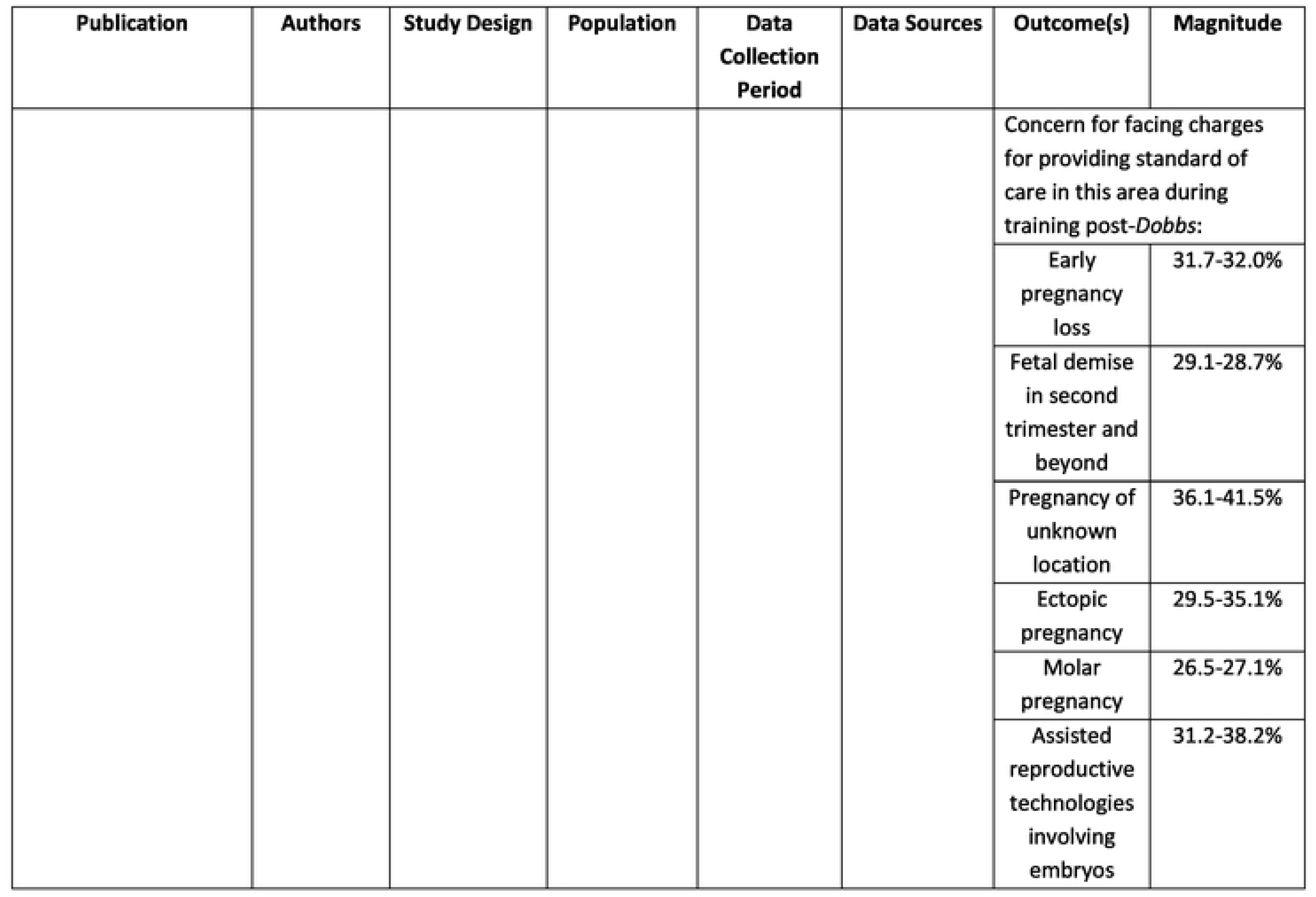
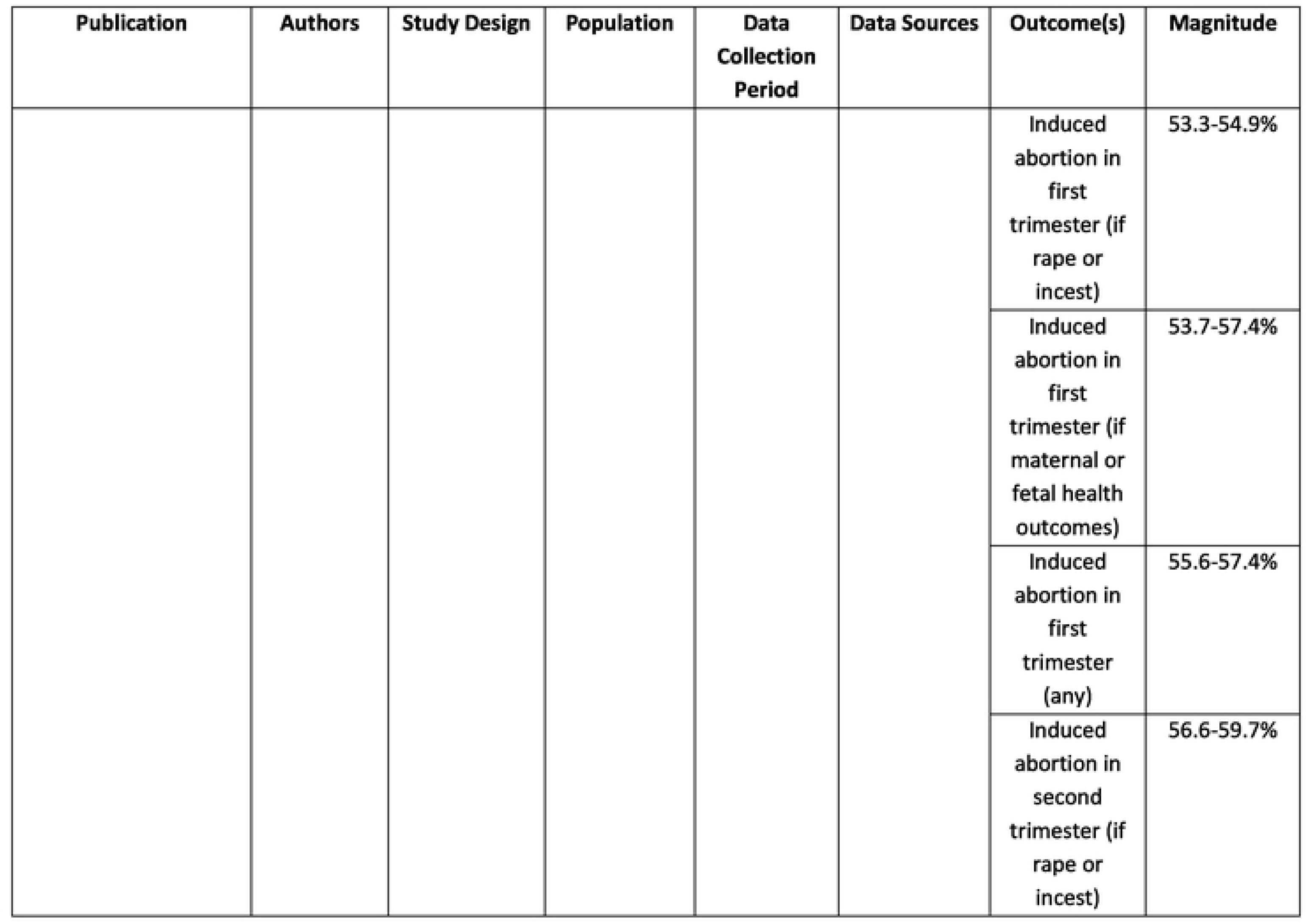
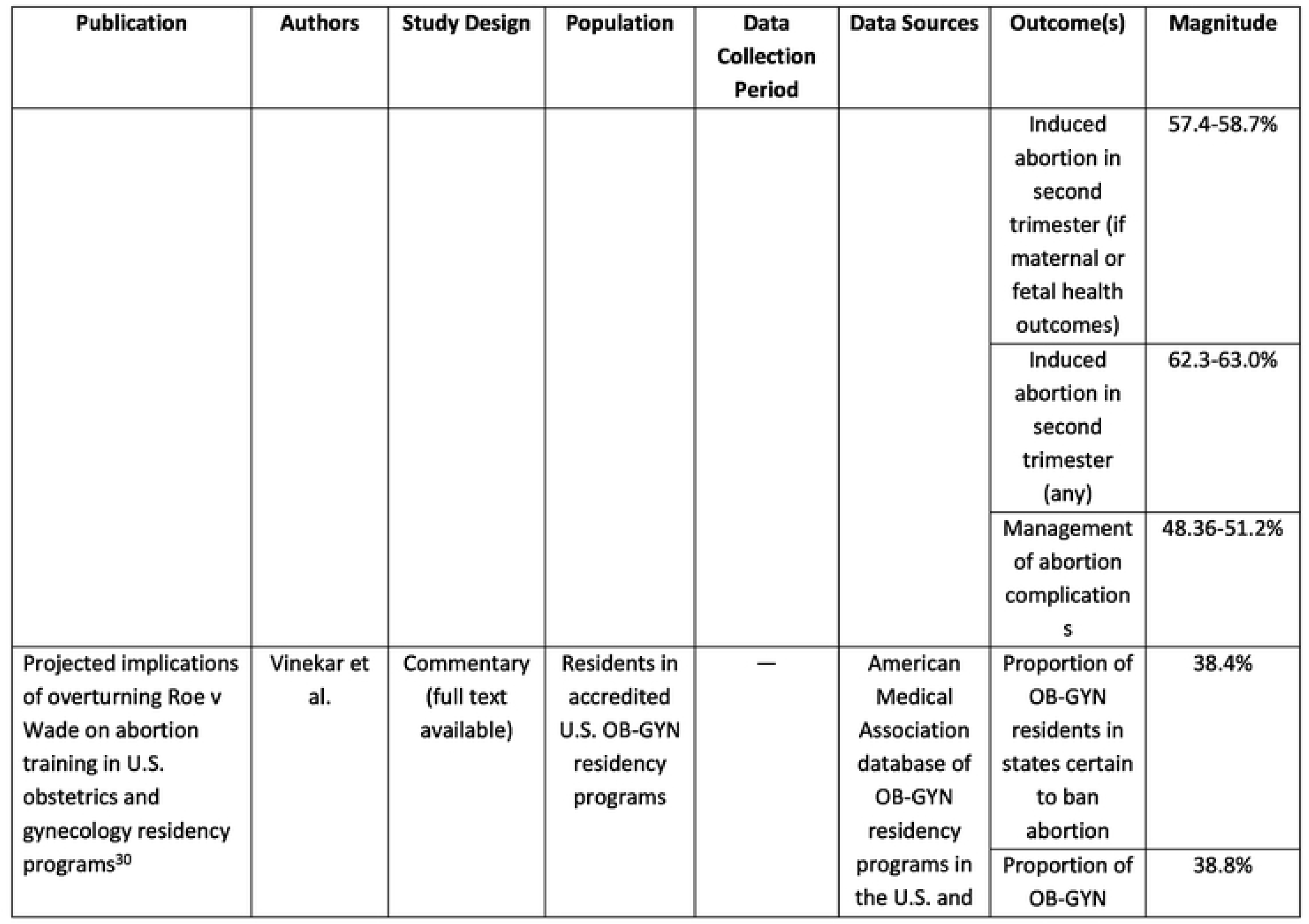
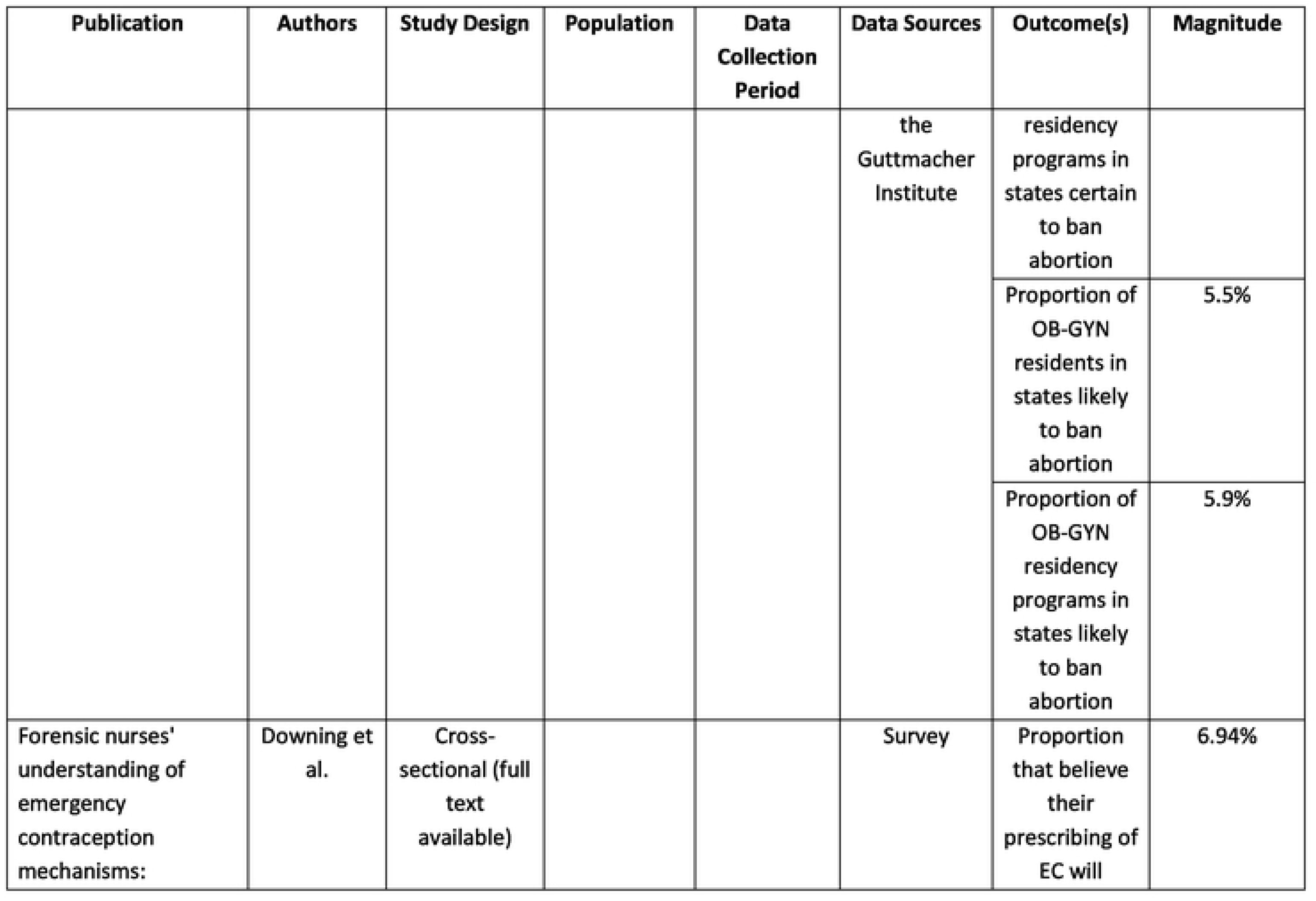
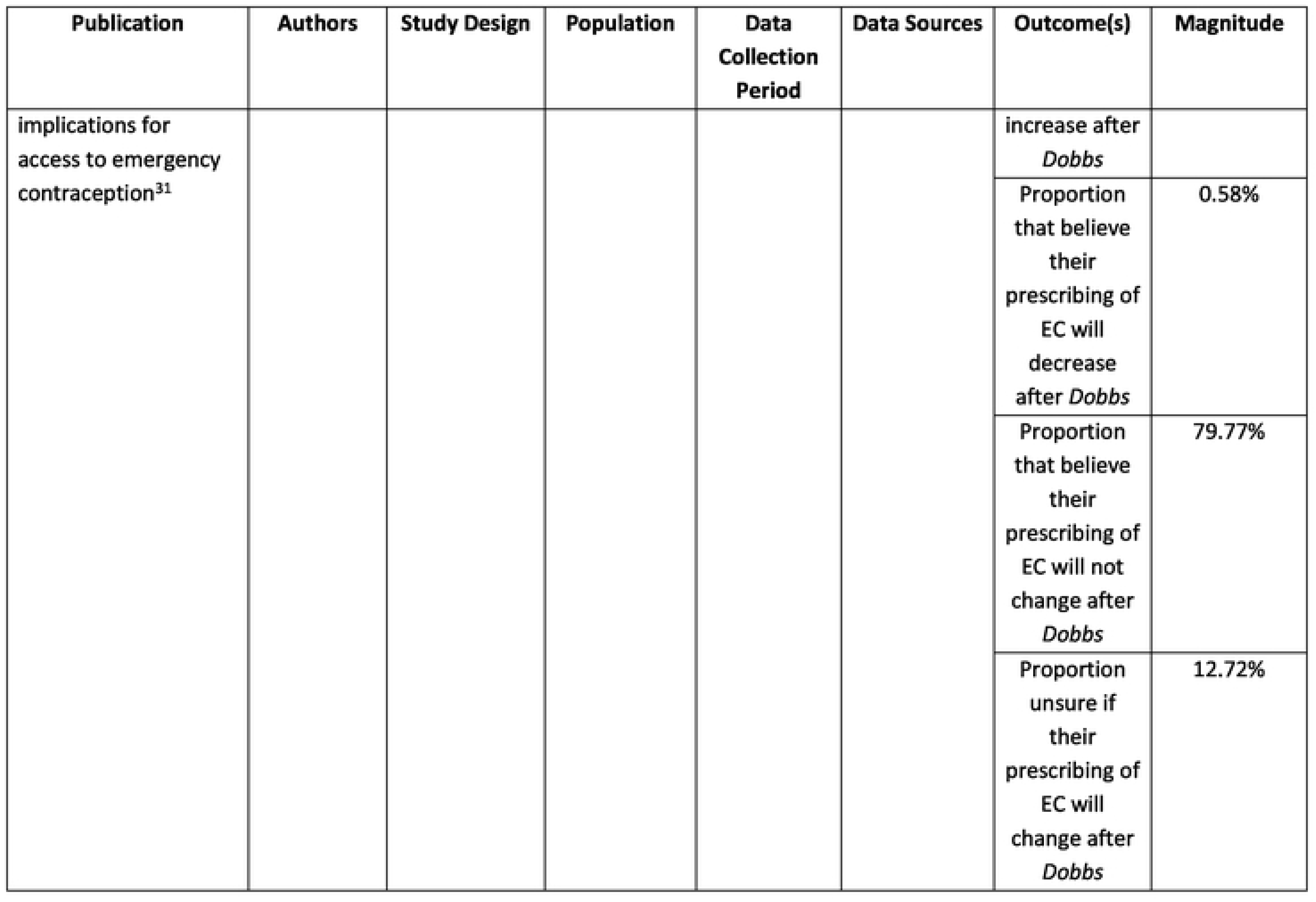
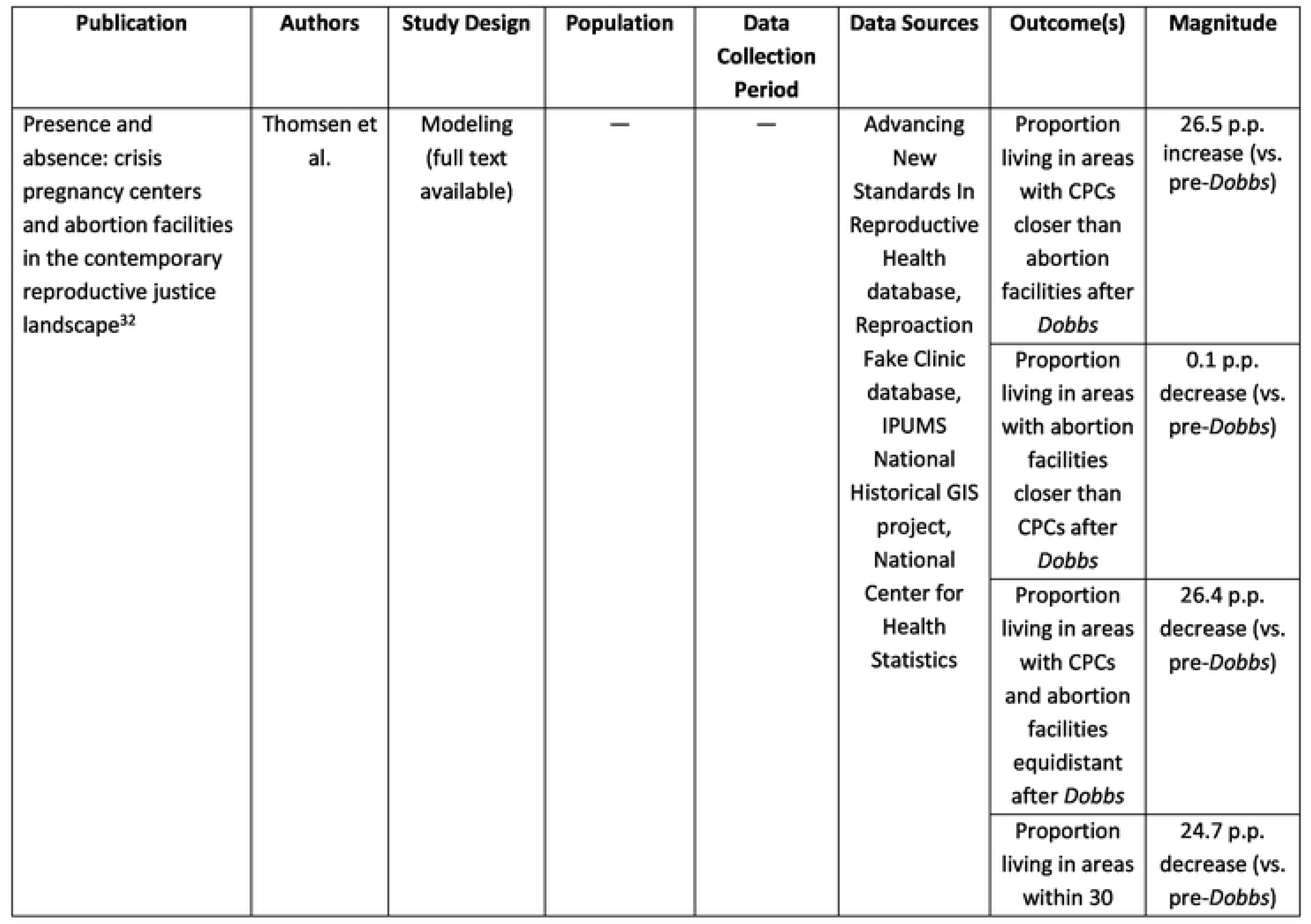
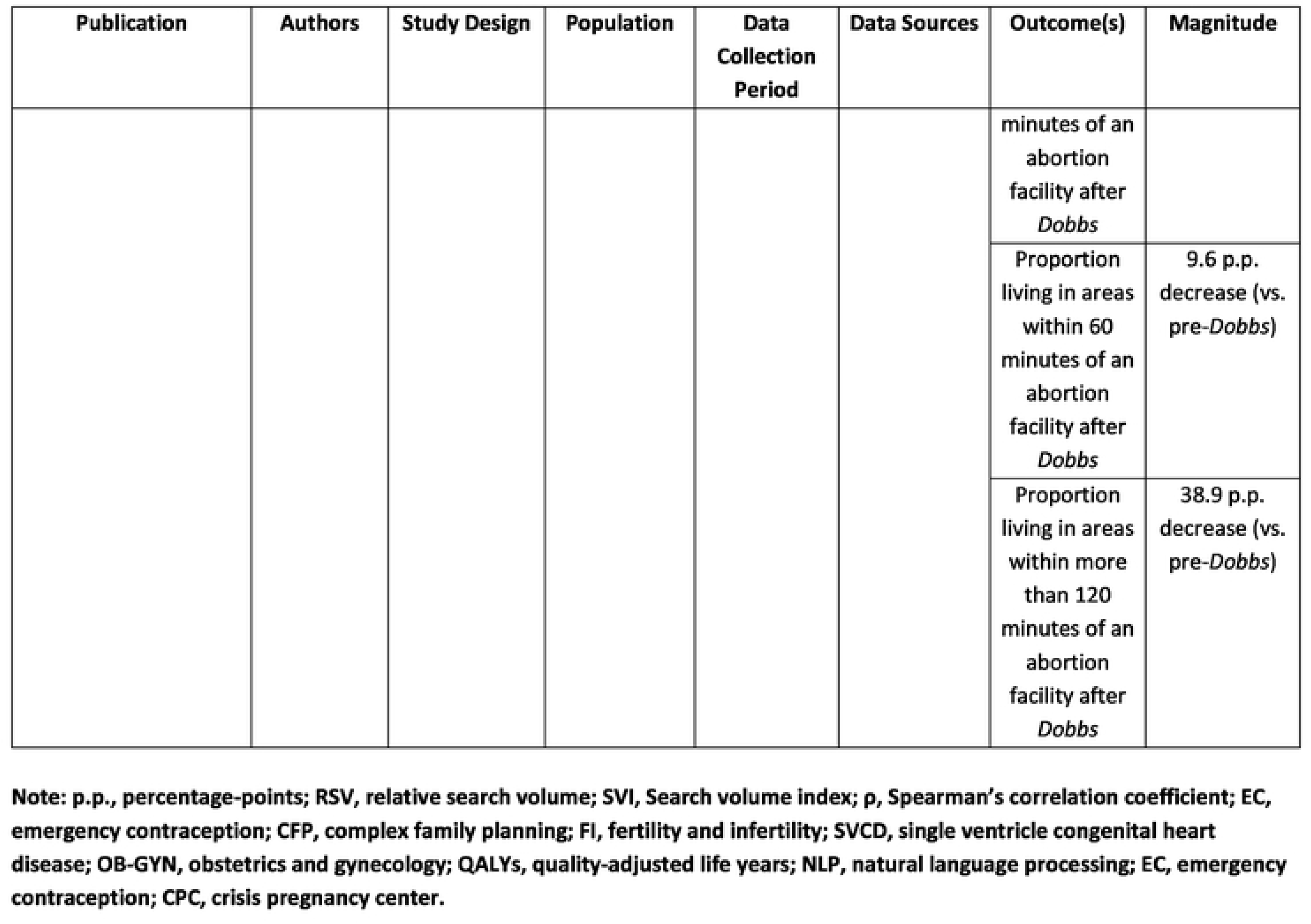
Study characteristics and major findings.

### 3.2 Contraception

Seven studies (38.9%) in our sample discussed contraception **(Table 2)**.^15–20, 31^ After *Dobbs*, we observed an increase in the demand for PCs. Notably, there was an increase in the number of Google searches for vasectomies immediately after *Dobbs* on June 24, 2022, as well as to a smaller extent immediately after the U.S. Supreme Court’s draft of *Dobbs v. Jackson* was leaked on May 2, 2022.^16–19^ The five states with the highest “vasectomy” search rates were Oklahoma, South Dakota, Idaho, New Mexico, and Hawaii.^19^ This is consistent with the increase of vasectomy requests, consultations, and procedures actually performed, identified using vasectomy billing data.^15^ Men who are younger (especially less than 30 years of age) and do not have children were more likely to seek vasectomy consultations after *Dobbs* was announced.^15^ Interestingly, the demand for vasectomies was inversely proportional to the ratio of urologists to adult men in states, therefore, *Dobbs* is expected to further strain the urological workforce in these states and increase delays.^16^ Similarly, there was an increase in the number of Google searches for “tubal ligation” after *Dobbs*, albeit to a lesser extent than for vasectomies.^18, 19^ The Northern and Southwestern regions of the U.S. had the highest surge in Google searches for vasectomies, whereas the Midwestern regions had the highest surge in tubal ligation searches.^18^

Similarly, the demand for ECs also surged after *Dobbs* **(Table 2)**. The number of Google searches for “morning after pill” rose by approximately 8-fold after *Dobbs* and was most pronounced in Idaho, District of Columbia, South Dakota, Oklahoma, and North Dakota.^19^ Maintaining access to ECs after *Dobbs* is vital to mitigate a wide array of adverse health outcomes, e.g., a modeling study predicted that maintaining access to ECs after *Dobbs* for a theoretical cohort of 750,000 patients capable of pregnancy was associated with a reduction in 41,052 abortions, 11,168 miscarriages, 1,611 cases of preeclampsia, 3,839 preterm births, 4 maternal deaths, 83 neonatal deaths, and 34 neurodevelopmental delays; in addition, it was associated with an additional 13,634 quality-adjusted life-years (QALYs) and US$541,716,923 in healthcare expenditure savings.^20^ Most forensics nurses believed that *Dobbs* would not affect prescribing of ECs (79.77%), although a minority believed that EC prescribing would increase (6.94%) and an even smaller proportion believed that this would decrease (0.58%).^31^ Several nurses cited concerns about current or legal restrictions around EC prescribing after *Dobbs* (e.g., fear of prosecution).^31^

Additionally, the demand for other contraceptives increased after *Dobbs* **(Table 2)**. The number of Google searches for “IUD”, “birth control pill”, and “condom” similarly spiked after *Dobbs*, albeit to a lower extent than PCs and ECs.^19^ Google searches for “IUD” were most pronounced in Utah, District of Columbia, Montana, Colorado, Minnesota, whereas Google searches for “condom” were most frequent in Delaware, New York, New Jersey, Connecticut, Mississippi.^19^ the modest spike in Google searches for “birth control pills” was suggested by the authors to be due to possible factors such as a lack of awareness, concerns about efficacy or side effects, cost-related barriers to access, or lack of convenience.^19^

### 3.3 Medications

Two studies (11.1%) in our sample discussed the impact of *Dobbs* on medications other than contraceptives **(Table 2)**.^22, 26^ One study found that requests for self-managed abortion medications via Aid Access, a telemedicine nonprofit organization that enables individuals to order abortion medications via mail,^33^ rose from 82.6 to 137.1 mean daily requests after the leaked draft of *Dobbs* on May 2, 2022, followed by a further increase to 213.7 mean daily requests after *Dobbs* officially passed.^22^ The requests rose in every U.S. state, although states that implemented total bans experienced the largest increase.^22^ The number of requesters citing “current abortion restrictions” as a primary reason increase from 31.4% to 62.4% after *Dobbs*, which tended to be most pronounced in abortion-hostile states but were also prevalent in states in which state laws governing abortion did not immediately change after *Dobbs*.^22^ Patients seeking teratogenic medications after *Dobbs* also faced difficulties. Notably, one study found that approximately 1 in 17 people experienced unexpected barriers to accessing methotrexate after *Dobbs*, of which 21.7% were directly related to *Dobbs* (e.g., prescription delays or refusals citing pregnancy risks or concerns related to abortion).^26^

### 3.4 Travel as a barrier to abortion access

Four studies (22.2%) in our sample discussed travel-related barriers to accessing abortion clinics and other reproductive health services **(Table 2)**.^21–24^ These studies consistently found that state laws under *Dobbs* would compound existing travel-related barriers. A modeling study predicted that the number of women facing restricted access to both contraception and abortion facilities would significantly increase after *Dobbs*, rising from 11% (pre-*Dobbs*) to 46% (post-*Dobbs*), affecting approximately 1.6 million women across 34 U.S. states.^23^ Similarly, another modeling study found that, between January 2021 to September 2022, the mean surface travel time (e.g., car or public transport) to abortion facilities increased from 27.8 minutes (pre-*Dobbs*) to 100.4 minutes (post-*Dobbs*), resulting in approximately 33.3% of reproductive-age women living in a census tract more than 60 minutes from an abortion facility compared to only 14.6% before *Dobbs*.^21^ Marginalized or medically-underserved populations were disproportionately affected by these travel-related barriers to accessing abortion care, e.g., census tracts located more than 60 minutes from an abortion facility predominantly consisted of residents of racial/ethnic minorities with a lower mean income and without health insurance, a high school diploma, or internet.^21^ The rise in prevalence of reproductive-age females living in a census tract more than 60 minutes from an abortion facility was higher among Black (25.6 p.p. increase), Hispanic (21.7 p.p. increase), and American Indian or Alaskan Native (20.4 p.p. increase) compared to White reproductive-age females (18.0 p.p. increase); in contrast, this change was smaller for Asian (14.1 p.p. increase) and Native Hawaiian or Pacific Islander reproductive-age females (11.8 p.p. increase).^21^ Racial/ethnic disparities were also documented with regards to the relative proximity of crisis pregnancy centers (CPCs) and abortion facilities. A modeling study predicted that, after *Dobbs*, the ratio of CPCs to abortion facilities would rise from approximately 1:3 to 1:5, affecting Hispanic (29.6 p.p. increase) more than White patients (24.4 p.p. increase), followed by Black (23.6 p.p. increase), Native American or Alaskan Native (20.6 p.p. increase), and Asian or Pacific Islander (19.2 p.p. increase) patients.^32^ Another survey found that those living in abortion-restricted states were more likely to experience travel-related barriers to access, rely on financial assistance, pay out-of-pocket for abortion care, and face financial barriers.^24^

### 3.5 Public attitudes

Only one study (5.6%) of our study sample discussed public attitudes towards *Dobbs* **(Table 2)**.^25^ By conducting sentiment analysis on tweets related to abortion and reproductive healthcare, they observed a growing polarization after *Dobbs*, in line with their expectations.^25^ Contrary to their expectations, this polarization was driven by a small (0.17 p.p.) increase in the proportion of overall negative/unfavorable tweets towards abortion and *Roe v. Wade*, as well as a modest (4.71 p.p.) decrease in positive/favorable tweets.^25^ These changes were most pronounced for tweets about “Roe v. Wade”, for which there was a 10.8 p.p. increase and 5.63 p.p. decrease in negative and positive tweets, respectively, followed by tweets about “family planning”, for which there was a 5.35 p.p. increase and 3.28 p.p. decrease in negative and positive tweets, respectively.^25^ Pro-life tweets typically centered around personal religious belief or support for conservative policies extending pro-life movements.^25^ In contrast, pro-choice tweets typically expressed anger and dismay with the *Dobbs* decision, highlighted the need for abortion, demonstrated fear over losing access to contraception after *Dobbs*, and perceived *Dobbs* as violating fundamental human rights for people capable of pregnancy.^25^

### 3.6 Medical residency and fellowship programs

Three studies (16.7%) in our sample discussed the implications of *Dobbs* on medical trainees, residents, and fellows **(Table 2)**.^28–30^ One study examined a sample of 286 obstetrics and gynecology (OB-GYN) residency training programs across the US and found that 38.8% of programs are located in states certain to ban comprehensive abortion care training after *Dobbs*, while an additional 5.9% are located in states likely to ban this training.^30^ This is equivalent to 43.9% of OB-GYN residents, totaling 2,638 individuals, that are training in programs located in states certain or likely to ban abortion after *Dobbs*.^30^ Significant concerns were brought up relating to the impact of *Dobbs* on restricting healthcare providers’ scope of practice. Notably, approximately 35.1-41.8% and 38.5-40.8% of residents and fellows in OB-GYN-related training programs across the U.S. believed that they would have to stop providing standard-of-care for induced abortion during the first and second trimester, respectively, after *Dobbs*.^29^ Similarly, approximately 38.5-40.8% and 62.3-63.0% expressed fear over facing charges for providing the standard-of-care for induced abortion during the first and second trimester, respectively, after *Dobbs*.^29^ This fear of legal consequences for providing standard-of-care abortion services was prevalent among residents and fellows after *Dobbs*, as evidenced by the following percentages: 48.4-51.2% for clinical management of abortion complications, 31.7-32.0% for early pregnancy loss care, 26.5-35.1% for caring for ectopic or molar pregnancies, and 31.2-38.2% for providing care using assisted reproductive technologies involving embryos.^29^ Maternal-fetal medicine (MFM) fellowship programs in abortion-hostile states are more likely to be associated with fertility and infertility (FI) centers, while MFM fellowships in abortion-friendly states are more likely to be associated with complex family planning (CFP) fellowship.^28^ Further, MFM fellows in abortion-friendly states are more likely to be female, participate in pro-abortion advocacy, and placed a higher emphasis on abortion-related training in their fellowship training.^28^

## Discussion

To the best of our knowledge, this is the first scoping review that examined the clinical implications of *Dobbs* on patients and healthcare providers or trainees (e.g., medical residents and fellows). Our early findings contribute to better understanding how *Dobbs* has transformed the current landscape of abortion and reproductive healthcare, spanning various forms of contraception, access to medications and health services, social and environmental barriers (e.g., travel- and cost-related barriers to access), public attitudes, and medical training programs. In the nascent stages of the post-*Dobbs* era, our findings offer valuable insights into further avenues for clinical and public health research to characterize the longer-term consequences of *Dobbs* on people with the capacity for pregnancy, as well as informing policy priorities and interventions that need to be taken to safeguard patient outcomes and abortion providers’ practices.

Notably, studies^15–20, 31^ frequently described the impact of *Dobbs* on demand for, and access to, PCs (e.g., vasectomies and tubal ligation), ECs, and other forms of contraception (e.g., IUD, birth control pill, condoms, etc.). The number of Google searches for contraception spiked after *Dobbs*, corroborating with an increase in the number of consultations and actual vasectomies performed as per billing data,^15^ demonstrating a greater interest and demand for contraception after *Dobbs*. This emphasizes the need for widespread access to contraception, reliable sources of information to guide decision-making (e.g., trusted online websites or improving access to healthcare professionals to guide abortion care, reproductive health services, or complex family planning), and active efforts to combat misinformation related to contraception. Further, we consistently observed that demand for, and access to, contraception after *Dobbs* was relatively lower among states with more restrictive abortion laws.^16, 18, 19^ This is concerning given the disproportionately higher number of contraceptive deserts in abortion-hostile states, underscoring the importance of redoubling efforts to increase equitable access to contraception.^34–36^ Of further concern, we observed that patients faced new barriers to other medications such as methotrexate after *Dobbs* due to providers’ hesitancy or refusal to prescribe them since they are capable of being teratogenic.^26^ This foreshadows the rising collateral impact and encroachment of *Dobbs* on other fields such as oncology or rheumatology, calling for cross-specialty and collective efforts in the healthcare system to address this issue.

Further, travel and financial concerns were frequently cited as barriers to accessing abortion in the post-*Dobbs* landscape.^21–24^ This has been extensively documented in the existing literature, complicating access to proper care due to the geographical inaccessibility of abortion facilities,^10^ legal restrictions (e.g., requiring cross-state travel to access reproductive health services),^37^ time constraints,^10^ cost burdens for those need to take time off work and find childcare services,^38^ emotional distress,^39^ among other mechanisms.^40^ These barriers are most pronounced for historically marginalized and medically-underserved communities, such as those who are Black, Latinx, uninsured, undocumented, low-income, and others, who are also the populations who tend to face the most health complications and adverse health outcomes from an abortion.^41^ Preliminary evidence shows that *Dobbs* is likely to expand and compound these barriers and disparities,^21–24^ emphasizing the need for cross-sectoral, multi-level partnerships to address these social and environmental barriers to abortion care after *Dobbs*.^36^

Finally, studies^28–30^ frequently discussed medical trainees’ and healthcare providers’ concerns about the impact of *Dobbs* on their scope of practice, particularly since a large proportion of OB-GYN-related residency and training programs are located in abortion-hostile states. Fellows frequently cited concerns over no longer being able to legally provide standard-of-care services for patients in need of abortions and similar reproductive health services, as well as a fear of legal prosecution and charges for offering these services amidst the new restrictions under *Dobbs*.^29^ It is clear that *Dobbs* has already severely limited medical trainees’ abilities to receive training for, and healthcare providers’ abilities to provide, the full spectrum of reproductive health services in abortion-hostile states. There is an urgent need for further advocacy from healthcare organizations at the regional, state, and national levels to safeguard and expand access to reproductive healthcare training and provision.

There are several limitations to our study. Firstly, we conducted a scoping review less than a year after *Dobbs* passed with the aim of conducting an exploratory analysis and serving a hypothesis-generating foundation for further studies. As such, our data sources were limited and our findings are not intended to be representative of the general U.S. population. Further, we did not attempt to quantitatively synthesize the proportions and rates overviewed in our study, thus, a meta-analysis may be warranted. More research is warranted to comprehensively describe the implications of *Dobbs* on patients and providers at a national level with a longer follow up period. Further, one-third of our articles reviewed were only available in their abstract form, thus, our findings are not final and may be subject to change once the full-text articles are made available in the future. Secondly, our screening process involved four reviewers, which may have introduced inconsistencies. Although we achieved a relatively high kappa score (0.82) and attempted to mitigate these inconsistencies by training all reviewers (e.g., screening a subset of articles together to help achieve a more consistent reasoning process), this may have still affected the reliability of our screening and findings. Finally, we excluded qualitative studies as we wanted to focus our analyses on observational studies, therefore, a future review that reviews qualitative studies and identify key themes related to patients’ and providers’ lived experiences after *Dobbs* may be appropriate.

## Conclusion

The clinical impact of *Dobbs v. Jackson* on patients’ health outcomes and access to health services, as well as providers’ abilities to continue providing the full spectrum of abortion and reproductive health services, is substantial. Further actions and research are needed from multiple spheres of action— healthcare providers, policymakers, legislators, public health agencies, and the public—to describe the consequences of *Dobbs* on the healthcare system and advocate for continued access to reproductive health services in the post-*Dobbs* landscape.

## Data Availability

All relevant data are within the manuscript and its Supporting Information files.

## Supporting information

**Table S1. PRISMA-ScR checklist.**

(DOCX)

**Table S2. Systematic review search strategy (restricted between 2022-2023)**

(DOCX)

## Author contributions

**Conceptualization:** David T. Zhu

**Data curation:** David T. Zhu, Lucy Zhao, Tala Alzoubi, Novera Shenin, Teerkasha Baskaran, Julia Tikhonov, Catherine Wang

**Formal analysis:** David T. Zhu, Lucy Zhao, Tala Alzoubi, Novera Shenin, Teerkasha Baskaran, Julia Tikhonov, Catherine Wang

**Investigation:** David T. Zhu, Lucy Zhao, Tala Alzoubi, Novera Shenin, Teerkasha Baskaran, Julia Tikhonov, Catherine Wang

**Methodology:** David T. Zhu

**Project administration:** David T. Zhu

**Supervision:** David T. Zhu

**Writing — original draft:** David T. Zhu

**Writing — review & editing:** David T. Zhu, Lucy Zhao, Tala Alzoubi, Novera Shenin, Teerkasha Baskaran, Julia Tikhonov, Catherine Wang

## Acknowledgements

Funding: No funding was received for this work.

## References

1. Supreme Court of the United States. Dobbs, state health officer of the Mississippi Department of Health, et al. v. Jackson Women’s Health Organization et al. June 24, 2022. https://www.supremecourt.gov/opinions/21pdf/19-1392_6j37.pdf. Date accessed: June 27, 2023.

2. Kaufman, R, Brown, R, Coral, CM, Jacob, J, Onyango, M, Thomasen, K. Global impacts of Dobbs v. Jackson Women’s Health Organization and abortion regression in the United States. Sex Reprod Health Matters. 2022;30(1):2135574. https://doi.org/10.1080/26410397.2022.2135574 PMID: 36383177

3. Guttmacher Institute. Interactive map: US abortion policies and access after Roe. July 5, 2023. https://states.guttmacher.org/policies/. Date accessed: June 27, 2023.

4. Raymond, EG, Grimes, D. The comparative safety of legal induced abortion and childbirth in the United States. Obstet Gynecol. 2012;119:215–219. https://doi.org/10.1097/AOG.0b013e31823fe923 PMID: 22270271

5. Guttmacher Institute. Abortion worldwide 2017: unevent progress and unequal access. 2017. https://www.guttmacher.org/sites/default/files/report_pdf/abortion-worldwide-2017.pdf. Date accessed: June 27, 2023.

6. Stevenson, AJ. The pregnancy-related mortality impact of a total abortion ban in the United States: a research note on increased deaths due to remaining pregnant. Demography. 2021;58(6):2019–2028. https://doi.org/10.1215/00703370-9585908 PMID: 34693444

7. Kozhimannil, KB, Hassan, A, Hardeman, RR. Abortion access as a racial justice issue. NEJM. 2022;387:1537–1539. https://doi.org/10.1056/NEJMp2209737 PMID: 36069823

8. Dehlendorf, C, Weitz, T. Access to abortion services: a neglected health disparity. JHCPU. 2011;22(2):415–421. https://doi.org/10.1353/hpu.2011.0064 PMID: 21551921

9. Redd, SK, Rice, WS, Aswani, MS, Blake, S, Julian, Z, Sen, B, et al. Racial/ethnic and educational inequities in restrictive abortion policy variation and adverse birth outcomes in the United States. BMC Health Serv Res. 2021;21:1139. https://doi.org/10.1186/s12913-021-07165-x PMID: 34686197

10. Bearak, JM, Lagasse, K, Jones, RK. Disparities and change over time in distance women would need to travel to have an abortion in the USA: a spatial analysis. Lancet Pub Health. 2017;2(11):e493–e500. https://doi.org/10.1016/S2468-2667(17)30158-5 PMID: 29253373

11. Tobin-Tyler, E. A grim new reality — intimate-partner violence after Dobbs and Bruen. NEJM. 2022;387:1247–1249. https://doi.org/10.1056/NEJMp2209696 PMID: 36193948

12. World Health Organization. Packages of interventions for family planning, safe abortion care, maternal, newborn and child health. 2010. https://apps.who.int/iris/handle/10665/70428. Date accessed: June 27, 2023.

13. Arksey, H, O’Malley, L. Scoping studies: towards a methodological framework. Int J Soc Res Methodol. 2005;19–32. https://doi.org/10.1080/1364557032000119616

14. Tricco, AC, Lillie, E, Zarin, W, O’Brien, KK, Colquhoun, H, Levac, D, et al. PRISMA extension for scoping reviews (PRISMA-ScR): checklist and explanation. Ann Intern Med. 2018,169(7):467-473. https://doi.org/10.7326/M18-0850 PMID: 30178033

15. Bole, R, Lundy, SD, Pei, E, Bajic, P, Parekh, N, Vij, SC. Rising vasectomy volume following reversal of federal protections for abortion rights in the United States. Nature. 2023. https://doi.org/10.1038/s41443-023-00672-x PMID: 36788351

16. Patel, RD, Loloi, J, Labagnara, K, Watts, KL. Search trends signal increased vasectomy interest in states with sparsity of urologists after overrule of Roe vs. Wade. 2022. https://doi.org/10.1097/JU.0000000000002901 PMID: 36082550

17. Sellke, N, Tay, K, Sun, HH, Tatem, A, Loeb, A, Thirumavalavan, N. The unprecedented increase in Google searches for “vasectomy” after the reversal of Roe vs. Wade. Fertil Sterill. 2022;118(6):1186–1188. https://doi.org/10.1016/j.fertnstert.2022.08.859 PMID: 36180257

18. Ghomeshi, A, Diaz, P, Henry, V, Ramasamy, R, Masterson, TA. The interest in permanent contraception peaked following the leaked Supreme Court majority opinion of Roe vs. Wade: a cross-sectional Google Trends analysis. Cureus. 2022;14(10):e30582. https://doi.org/10.7759/cureus.30582 PMID: 36420253

19. Datta, PK, Chowdhury, SR, Aravindan, A, Nath, S, Sen, P. Looking for a silver lining to the dark cloud: a Google Trends analysis of contraceptive interest in the United States post Roe vs. Wade verdict. Cureus. 2022;14(7):e27012. https://doi.org/10.7759/cureus.27012 PMID: 35989835

20. Dzubay, SK, Doshi, U, Chaiken, SR, Arora, M, Caughey, AB. Impact of banning emergency contraception in states with abortion bans: a cost-effectiveness analysis. Am J Obstet Gynecol. 2023;223(1):S734. Accessed: June 27, 2023. https://doi.org/10.1016/j.ajog.2022.11.1225

21. Rader, B, Upadhyay, UD, Sehgal, NKR, Reis, BY, Brownstein, JS, Hswen, Y. Estimated travel time and spatial access to abortion facilities in the US before and after the Dobbs v Jackson Women’s Health decision. JAMA. 2022;328(20):2041–2047. https://doi.org/10.1001/jama.2022.20424 PMID: 36318194

22. Aiken, ARA, Starling, JE, Scott, JG, Gomperts, R. Requests for self-managed medication abortion provided using online telemedicine in 30 US states before and after the Dobbs v Jackson Women’s Health Organization decision. JAMA. 2022;328(17):1768–1770. https://doi.org/10.1001/jama.2022.18865 PMID: 36318139

23. Rodriguez, MI, Meath, THA, Watson, K, Daly, A, Myers, C, McConnell, KJ. Predicted changes in travel distance for abortion among counties with low rates of effective contraceptive use following Dobbs v Jackson. Am J Obstet Gynecol. 2023;228(6):752–753. https://doi.org/10.1016/j.ajog.2023.01.032 PMID: 36738910

24. Jones, RK, Chiu, DW. Characteristics of people obtaining abortions in states likely to ban it: findings from a 2021-2022 national study. https://doi.org/10.1016/j.contraception.2022.09.020. Accessed: June 27, 2023.

25. Mane, H, Yue, X, Yu, W, Doig, AC, Wei, H, et al. Examination of the public’s reaction on Twitter to the over-turning of Roe v Wade and abortion bans. Healthcare. 2022;10(12):2390. https://doi.org/10.1016/j.ajog.2023.01.032 PMID: 36738910

26. Wipfler, K, Cornish, A, Schumacher, R, Shaw, Y, Katz, P, Michaud, K. Impact on access to methotrexate in the post-Roe era. Arthritis Rheumatol. 2022;74(suppl 9). Contraception. 2022;116:72. https://acrabstracts.org/abstract/impact-on-access-to-methotrexate-in-the-post-roe-era/. Accessed: June 27, 2023.

27. Miller, HE, Henkel, A, Zhang, J, Leonard, SA, Quirin, AP, Maskaia, SA, et al. Abortion restriction impact on burden of neonatal single ventricle congenital heart disease: a decision-analytic model. Am J Obstet Gynecol. 2023;228(1):S483. https://doi.org/10.1016/j.ajog.2022.11.834. Accessed: June 27, 2023.

28. Cheng, C, Byrne, JJ, Hernandez, Michalek, JE, Pierce, CM, Martinez, M, et al. Fellow perspectives of abortion-related training in maternal-fetal medicine fellowship: regional differences in a post-Roe world. Am J Obstet Gynecol. 2023;228(1):S106–S107. https://doi.org/10.1016/j.ajog.2022.11.220. Accessed: June 27, 2023.

29. Meriwether, KV, Krashin, JW, Kim-Fine, S, Avlove, T, Dale, L, Orejuela, FJ, et al. Trainee opinions regarding the effect of the Dobbs v. Jackson women’s health organization Supreme Court decision on obstetrics and gynecology training. Am J Obstet Gynecol. 2023;228(3):S816–S817. https://doi.org/10.1016/j.ajog.2022.12.045. Accessed: June 27, 2023.

30. Vinekar, K, Karlapudi, A, Nathan, L, Turk, JK, Rible, R, Steinauer, J. Projected implications of overturning Roe v Wade on abortion training in U.S. obstetrics and gynecology residency programs. Obstet Gynecol. 2022;140(2):146–149. https://doi.org/10.1097/AOG.0000000000004832 PMID: 35852261

31. Downing, NR, Avshman, E, Valentine, JL, Johnson, LM, Chapa, H. Forensic nurses’ understanding of emergency contraception mechanisms: implications for access to emergency contraception. J Forensic Nurs. 2023. https://doi.org/10.1097/JFN.0000000000000430 PMID: 36917678

32. Thomsen, C, Levitt, Z, Gernon, C, Spencer, P. Presence and absence: crisis pregnancy centers and abortion facilities in the contemporary reproductive justice landscape. Hum Geogr J. 2022;16(1):64–74. https://doi.org/10.1177/19427786221109959

33. Aid Access. https://aidaccess.org/en/. Date accessed: June 27, 2023.

34. Kreitzer, RJ, Smith, CW, Kane, KA, Saunders, TM. Affordable but inaccessible? Contraception deserts in the US states. J Health Polit Policy Law. 2021;46(2):277–304. https://doi.org/10.1215/03616878-8802186 PMID: 32955562

35. Salganicoff, A, Ranji, U. A focus on contraception in the wake of Dobbs. WHI. June 13, 2023. https://www.kff.org/womens-health-policy/perspective/a-focus-on-contraception-in-the-wake-of-dobbs/. Date accessed: June 27, 2023.

36. Zhu, DT. Cross-sectoral community and civic engagement after Dobbs v. Jackson. Lancet Reg Health Am. 2023;22:100514. https://doi.org/10.1016/j.lana.2023.100514 PMID: 37250688

37. Borgmann, CE, Jones, BS. Legal issues in the provision of medical abortion. Am J Obstet Gynecol. 2000;183(2):S84–S94. https://doi.org/10.1067/mob.2000.108229 PMID: 10944373

38. Barr-Walker, JB, Jayaweera, RT, Ramirez, AM, Gerdts, C. Experiences of women who travel for abortion: A mixed methods systematic review. PLoS ONE. 2019;14(4): e0209991. https://doi.org/10.1371/journal.pone.0209991 PMID: 30964860

39. Kimport, K, Rasidjan, MP. Exploring the emotional costs of abortion travel in the United States due to legal restriction. Contracept. 2023;120:109956. https://doi.org/

40. Doran, F, Nancarrow, S. Barriers and facilitators of access to first-trimester abortion services for women in the developed world: a systematic review. BMJ Sex Reprod Health. 2015;41;3:170–180. https://doi.org/10.1371/journal.pone.0209991 PMID: 30964860

41. Studnicki, J, Fisher, JW, Sherley, JL. Perceiving and addressing the pervasive racial disparity in abortion. Health Serv Res Manag. 2020;7:1–4. https://doi.org/10.1177/2333392820949743 PMID: 32875006

